# Type 2 Deiodinase in Cancer-Associated Fibroblasts: A Potential Therapeutic Target in Many Types of Cancer

**DOI:** 10.64898/2026.09.14.26362848

**Authors:** Mary Grace Carroll, Noha Mukhtar, Jacob Calhoun, Breaunna Garza, Matthew A. Loberg, Hua-Chang Chen, Quanhu Sheng, Alyse Staley, Tatiana L. Fonseca, Antonio C. Bianco, Matthew D. Ringel, Pamela Brock, Andrew J. Gunderson, Ahmad A. Tarhini, W. Douglas Cress, Michael J. Cavnar, Stephen Edge, Varinder Kaur, George J. Weiner, Janice L. Farlow, Robert Dood, Roman Groisberg, Abdul Rafeh Naquash, Craig Shriver, Dinesh Pal Mudaranthakam, Jose A. Oliveras, Robert J Rounbehler, Michelle L. Churchman, Vivian L. Weiss, Rebecca E. Schweppe, Nikita Pozdeyev, Bryan R Haugen

## Abstract

**Background:** Cancer-associated fibroblasts (CAF) are major regulators of the tumor microenvironment (TME), yet the pathways governing CAF function remain incompletely understood. Type 2 deiodinase (DIO2, D2), which converts thyroxine (T4) to the active thyroid hormone triiodothyronine (T3), has not been systematically investigated in CAF across different solid tumors.

**Methods:** We analyzed *DIO2* expression using single-cell RNA sequencing (scRNA-seq) from primary thyroid tumors, a large integrated scRNASeq thyroid cancer atlas, and publicly available scRNA-seq datasets representing six additional solid tumor types. Bulk RNA sequencing data from the Oncology Research Information Exchange Network (ORIEN) database were used to examine correlations among *DIO2*, CAF subtype markers, and overall survival (OS). A genetically engineered TPO-CreERT2/BrafV600E/Trp53-deficient (TBP) mouse model was evaluated for CAF-specific *Dio2* expression and pharmacologic inhibition of D2 activity. TBP mice were crossed with *Dio2*^-/-^ (Dio2 KO) mice to investigate the role of Dio2 in cancer development and progression.

**Results:** *DIO2* expression was highly restricted to CAF and was largely absent from epithelial-derived cancer, immune, and endothelial cells. Across thyroid cancer and six additional malignancies, *DIO2* was consistently detected in CAF, demonstrating that CAF-specific DIO2 expression is a conserved feature of multiple tumor types. Within thyroid cancer, *DIO2* was enriched in myofibroblastic CAF (myCAF) compared with inflammatory CAF (iCAF), and bulk transcriptomic analyses demonstrated stronger correlations between DIO2 expression and canonical myCAF markers than iCAF markers across numerous cancers. Elevated *DIO2* expression was associated with worse OS in pancreatic and bladder cancers, paralleling the adverse prognostic impact of increased myCAF abundance. In contrast, thyroid cancer demonstrated an inverse association attributable to loss of epithelial DIO2 expression during tumor dedifferentiation. The TBP mouse model recapitulated human disease, with *Dio2* expression restricted to CAF, robust stromal fibrosis, and measurable D2 enzymatic activity that was suppressed by cefuroxime and lenvatinib. Genetic loss of *Dio2* in the TME resulted in significantly reduced tumor growth.

**Conclusions**

DIO2 is a conserved marker of CAF, and myCAF, across multiple solid tumors and is associated with adverse clinical outcomes in some cancers. The TBP mouse provides a robust preclinical model to define the mechanistic role of DIO2 in CAF biology and to evaluate DIO2-directed therapeutic strategies targeting the tumor microenvironment.

## Introduction

Cancer-associated fibroblasts (CAF) are a major cellular component of the tumor microenvironment (TME) and play a central role in tumor progression, metastasis, and therapeutic resistance. Once considered a relatively uniform stromal population, CAF are recognized as a highly heterogeneous, extremely plastic population comprising multiple functionally distinct subtypes, including myofibroblastic CAF (myCAF) and inflammatory CAF (iCAF) (1, 2). MyCAF are typically characterized by extracellular matrix (ECM) production and contractile features, whereas iCAF secrete cytokines and chemokines that modulate immune responses. Across multiple tumor types, increased abundance of CAF, particularly myCAF, has been associated with worse clinical outcomes, likely due to their roles in matrix remodeling, immune exclusion, and promotion of tumor cell survival and invasion (2, 3). Despite these advances, key metabolic and signaling pathways that define CAF function across cancers remain incompletely understood.

Thyroid hormone signaling has emerged as an important regulator of cancer biology, influencing proliferation, differentiation, metabolism, and angiogenesis. The active thyroid hormone, triiodothyronine (T3), exerts its effects primarily through nuclear thyroid hormone receptors (THRA and THRB), regulating transcriptional programs involved in cell growth and metabolic activity. Both tumor-promoting and tumor-suppressive roles of thyroid hormone signaling have been described, depending on tumor type cellular context, and patient age (4–7). In several solid tumors, thyroid hormones have been shown to enhance cancer cell proliferation, mitochondrial function, and angiogenesis, while in other contexts it may promote differentiation and restrain tumor aggressiveness. However, most studies have focused on tumor cell–intrinsic effects, with relatively little attention to how thyroid hormone signaling is spatially regulated within the TME.

Local regulation of thyroid hormone activity is controlled by the iodothyronine deiodinases, a family of enzymes that activate or inactivate thyroid hormone. Type 2 deiodinase (DIO2, D2) converts the prohormone thyroxine (T4) into active T3, whereas type 3 deiodinase (DIO3) inactivates T3 and T4, thereby limiting thyroid hormone signaling. These enzymes play critical roles in tissue-specific control of thyroid hormone action during development and in adult physiology. In cancer, dysregulated expression of deiodinases has been reported in tumor cells, with DIO3 frequently associated with dedifferentiation and aggressive disease phenotypes, and DIO2 implicated in promoting proliferation and metabolic activity in certain contexts (8, 9).

However, the cell-type–specific expression and function of deiodinases within the TME, particularly in stromal compartments such as CAF, remain poorly defined. De Stefano and colleagues recently published *DIO2* expression in thyroid cancer CAF using scRNASeq from publicly available data (10).

Crosstalk between stromal and tumor cells is a key determinant of tumor behavior, with CAF acting as important regulators of nutrient availability, redox balance, and paracrine signaling within the TME (11, 12). Given the central role of deiodinases in controlling intracellular thyroid hormone levels, it is plausible that stromal cells could serve as localized sources of active T3, thereby shaping signaling gradients within tumors. However, whether CAF contribute to thyroid hormone signaling in cancer, and how this may influence CAF phenotype and tumor progression, has not been systematically investigated.

In this study, we sought to define the expression and functional role of DIO2 in CAF across thyroid cancer and other solid tumors. Using single-cell and bulk transcriptomic analyses, as well as *in vivo* modeling, we identify *DIO2* expression in a subset of CAF, particularly myCAF, and propose that local T3 production within stromal cells may represent a previously unrecognized mechanism regulating the TME.

## METHODS

### Human and murine scRNAseq sample acquisition and preprocessing

The initial anaplastic thyroid cancer (ATC) tumor sample (CCDC6-RET fusion, c.1-124C>T TERT promoter mutation, TP53 pR280K mutation) was collected from a patient undergoing surgical resection of the tumor, through an IRB approved protocol (COMIRB 07-0562). The murine thyroid tumor sample was harvested from a female TBP mouse 20 weeks after injection with tamoxifen. Tumors were measured, weighed, and immediately washed in serum-free RPMI. After mincing to approximately 2mm pieces, the sample underwent digestion with Liberase TM for 30min at 37°C. Fetal bovine serum (FBS) was added to stop the reaction, and subsequent steps were performed on ice. The tumor digest was gently disrupted using a serological pipette, then the tube was left on ice until the solids settled. The supernatant was washed with RPMI media supplemented with 10% FBS. Next, red blood cells were lysed using 1X RBC Lysis Buffer (eBiosciences) with a 4-minute incubation at room temperature. Cells were washed again and passed through a 30 µm mesh filter (Celltrics). Filtered cells were then washed twice with phosphate-buffered saline (PBS) + 0.04% bovine serum albumin (BSA) before counting (ViCell) and resuspending to 1,000 cells/uL for sequencing. Single-cell suspensions were processed using a 10X Genomics Single Cell 3’ v2 library and Chromium system. Next-generation sequencing was performed on a Novo-Seq 6000 instrument (Illumina). Initial mapping to the hg38 (human) or mm10 (murine) reference genomes, barcode processing, unique molecular identifier (UMI) counting, and gene expression estimation were performed using Cell Ranger 7.0.0 software from 10x Genomics.

scRNAseq data from non-thyroid cancers were acquired from NCBI’s Gene Expression Omnibus. Raw UMI count matrices and metadata were downloaded from GEO datasets <u>GSE229413</u>, <u>GSE131907</u>, <u>GSE200997</u>, <u>GSE176031</u>, and <u>GSE180286</u> (13–18).

### scRNAseq analysis

Analyses were performed in R version 4.3.1 using the Seurat package version 4.4 (19). Doublets were discovered from each sample using the R package *DoubletFinder* (20). Quality control was performed by removing single cells with greater than 25% mitochondrial gene expression, fewer than 300 detected genes, or fewer than 500 unique molecular identifiers (UMI) counts.

Following quality control, gene expression counts were log-normalized, and the 2,000 most highly variable genes were identified and scaled. Non-thyroid cancer samples were integrated using Seurat’s anchor-based integration method to account for batch effects. To determine the optimal number of principal components (PCs), elbow and JackStraw plots were generated across 50 PCs. Candidate PC numbers were selected based on where variance stabilized across both plots, then evaluated using Louvain clustering. The smallest number of PCs that preserved cluster number and biological resolution was selected and subsequently used for UMAP dimensionality reduction. Cell populations were annotated using a panel of cell type-specific markers and the top 30 differentially expressed genes of each cluster. In thyroid cancer samples, the following markers were included to identify thyroid-derived cell populations: thyroid follicular cells – *NKX2-1, PAX8, FOXE1*; thyroid differentiation markers – *TG, TPO, DIO1, DIO2, SLC5A5*.

### Analysis of deiodinase expression in a thyroid cancer single-cell atlas

The single-cell atlas of thyroid cancer is composed of 81 samples from 7 published data sets spanning benign (normal and paratumor samples, n=20), PTC (n=39), and ATC (n=22) tumor samples (21). The atlas was constructed, and major stromal subclusters were identified.

Deiodinase expression was evaluated in stromal subclusters using *Seurat* functions *FeaturePlot*, *VlnPlot*, and *DotPlot*.

### Bulk RNAseq correlation and survival analyses

To investigate associations between CAF composition and DIO2 expression across tumor types in larger datasets, we queried the Oncology Research Information Exchange Network (ORIEN) database. Normalized mRNA levels were generated and provided to the investigator team by Aster Insights (https://www.oriencancer.org/the-orien-difference). Analyses were restricted to cancer types with >500 tumors. myCAF content was estimated using six markers (*FAP, POSTN, COL1A2, ACTA2, PDGFRA, MMP11*), and iCAF content was estimated using three markers (*CXCL12, DPT, APOD*). Correlations between CAF markers and *DIO2/DIO3* expressions were assessed across tumor types, and linear regression models were fit to estimate R^2^ values. To evaluate associations with overall survival (OS), Kaplan-Meier analysis was performed using median FAP or *DIO2* mRNA expression as a binary cutoff. Complementary univariate Cox proportional hazards (CoxPH) modeling of OS was conducted using CAF markers and *DIO2* as continuous variables, with hazard ratios reflecting a one-standard-deviation (SD) increase in each variable. KM and CoxPH analyses were performed using the survival and survminer packages in R version 4.5.1 (https://CRAN.R-project.org/package=survival).

### DIO2 expression in normal thyroid tissue, thyroid cancer subtypes, and thyroid cancer cell lines

Raw gene expression data (Affymetrix GeneChip Human Genome U133 Plus 2.0) for normal thyroid tissue, papillary thyroid cancer (PTC), poorly differentiated thyroid cancer (PDTC), and anaplastic thyroid cancer (ATC) were downloaded from Gene Expression Omnibus (22) and GSM2024835 (23). Thyroid cancer cell line gene expression data was acquired from our prior publication (24). To reduce batch effect, background subtraction, normalization and summarizing probe sets were done jointly for CEL files from all three sources.

### Animal studies

***T****PO-CreER^T2^/**B**raf^V600E/wt^/Tr**p**53*^Δ*ex2–10/*Δ*ex2–10*^ mice, referred to as TBP, were used for these studies (25, 26). At 6-8 weeks of age, TBP mice are treated with tamoxifen (0.1 mg/g intraperitoneally for 2 days) to induce tumors. Following tamoxifen administration, mice receive drinking water supplemented with 0.2 mg/L levothyroxine. At 20 weeks of age, mice undergo thyroid ultrasound every 2 weeks to track tumor formation. When tumor volumes were 25-75mm^3^, mice were randomized to vehicle (n=4) or cefuroxime (DR inhibitor, n=4) treatment groups. Mice received intraperitoneal injections daily for 15 days, with the cefuroxime-treated mice receiving 900 mg/kg/day (27). At harvest, tumors were collected and snap frozen for measurement of D2 activity. In a separate study, untreated TBP tumors were resected, formalin-fixed and paraffin-embedded (FFPE) for histology and immunohistochemistry.

B6.129-Dio2^tm1Vag^/J mice were purchased from The Jackson Laboratory (strain #018985) (28). To determine the role of Dio2 on tumor growth in the TBP mice, the TBP mice were crossed with *Dio2*^-/-^ (Dio2 KO) mice to eventually generate parental pairs of mice that were *TPO-CreER^T2^*/*Braf^V600E^*^/V600E^/*Trp53*^Δ*ex2–10/*Δ*ex2–10*^*/Dio2^+/-^*crossed with *TPO-CreER^T2^/Braf^WT/WT^/Trp53*^Δ*ex2–10/*Δ*ex2–10*^*/Dio2^+/-^*to generate experimental littermates (*TPO-CreER^T2^/Braf^V600E/WT^/Trp53*^Δ*ex2–10/*Δ*ex2–10*^ with WT or KO Dio2). Tumor induction in TBP-Dio2 animals at 6–8 weeks was performed by tamoxifen administration (0.16 mg/g in corn oil, intraperitoneal [i.p.] daily for 2 days). Thyroid hormone (levothyroxine; 0.2 mg/L in 0.75% ethanol) was given in drinking water following tumor induction. Beginning 10 weeks after tamoxifen induction, thyroid tumors were measured by ultrasound and monitored for 14 weeks in TBP-Dio2WT (n= 19) and TBP-Dio2KO (n= 16) littermate mice. Statistical significance was measured using a two-way ANOVA test.

### Deiodinase 2 activity assay

Thyroid tumor samples were homogenized and sonicated in buffer solution containing 20 mM dithiothreitol (DTT) and 0.25 M sucrose, as previously described (29). Enzyme activity was assayed by incubating 60 μg of protein with 0.2 nM [¹² I]-5′-T4 (Revvity, Boston, MA) as substrate for 4 h at 37 °C. All reactions contained 1 mM PTU to inhibit type 1 deiodinase activity and 20 mM DTT as a cofactor. ¹² I released was quantified using a γ-counter, and the activity was expressed as fmol/h/mg protein.

### Col1a1 immunohistochemistry

Five-micron sections of TBP tumors were cut and stained for Col1a1 to visualize CAF infiltrate and fibrosis (Cell Signaling #72026; 1:100 dilution). Standard procedures for IHC were used.

First, slides went through a standard staining protocol to deparaffinize and rehydrate the tissue. Then, antigen retrieval was performed, followed by blocking with 3% hydrogen peroxide and normal goat serum. SignalStain Diaminobenzene (DAB) Substrate (Cell Signaling) was used to visualize Col1a1, then the slides were counterstained with Hematoxylin. Slides were then imaged and scanned into the Aperio Digital Pathology system. After training the system, digital scanning produced areas of positive staining (% pixels) and intensity of staining (1+, 2+ or 3+).

## RESULTS

### DIO2 is expressed in CAF from a patient with ATC

To better understand the tumor microenvironment (TME) in advanced thyroid cancer, we performed scRNASeq on a patient who had surgically resected ATC. This female in her 60’s had an incidentally discovered left thyroid nodule. FNA was suggestive of undifferentiated thyroid cancer. Histopathological examination of the resected tissue revealed (ATC) with a small component of PTC and metastasis to 3 central neck lymph nodes. Molecular testing revealed a *CCDC6-RET* fusion, a *TERT* c.1-124C>T mutation and a TP53 p.R280K (c.839G>A) mutation.

For scRNASeq, 5612 cells were sequenced with an average of 27,163 UMI/cell and an average of 4335 genes per cell sequenced. There were 16 distinct clusters identified. Figure 1 shows identification of clusters using markers for thyrocyte-derived cells and epithelial cells (Fig 1A), thyrocyte differentiation markers (Fig 1B), myeloid markers (Fig 1C), T cell markers (Fig 1D) and CAF markers (Fig 1E). Fig 2A labels the four major clusters (myeloid cells, thyroid cancer cells, T cells and CAF). Interestingly, a thyroid differentiation marker, *DIO2*, was identified only in the CAF cluster in the UMAP plot (Fig 2A), which is also shown in the Violin plot (Fig 2B) and Dot plot (Fig 2C) that *DIO1* or *DIO3* are not expressed in any of the cell clusters. The primary role for *DIO2* in the cell is to convert the prohormone T4 into the active hormone T3, through the thyroid hormone nuclear receptors (THR) THRA and THRB. Fig 2D shows that *THRA* is the predominantly expressed THR, with the highest expression in approximately 40% of the CAF and lower levels of expression in the thyroid cancer cells. These data may suggest that there is local conversion of T4 to T3 in CAF from this ATC patient, and that the CAF themselves may be a primary target of T3 in the TME.

**Figure 1.**
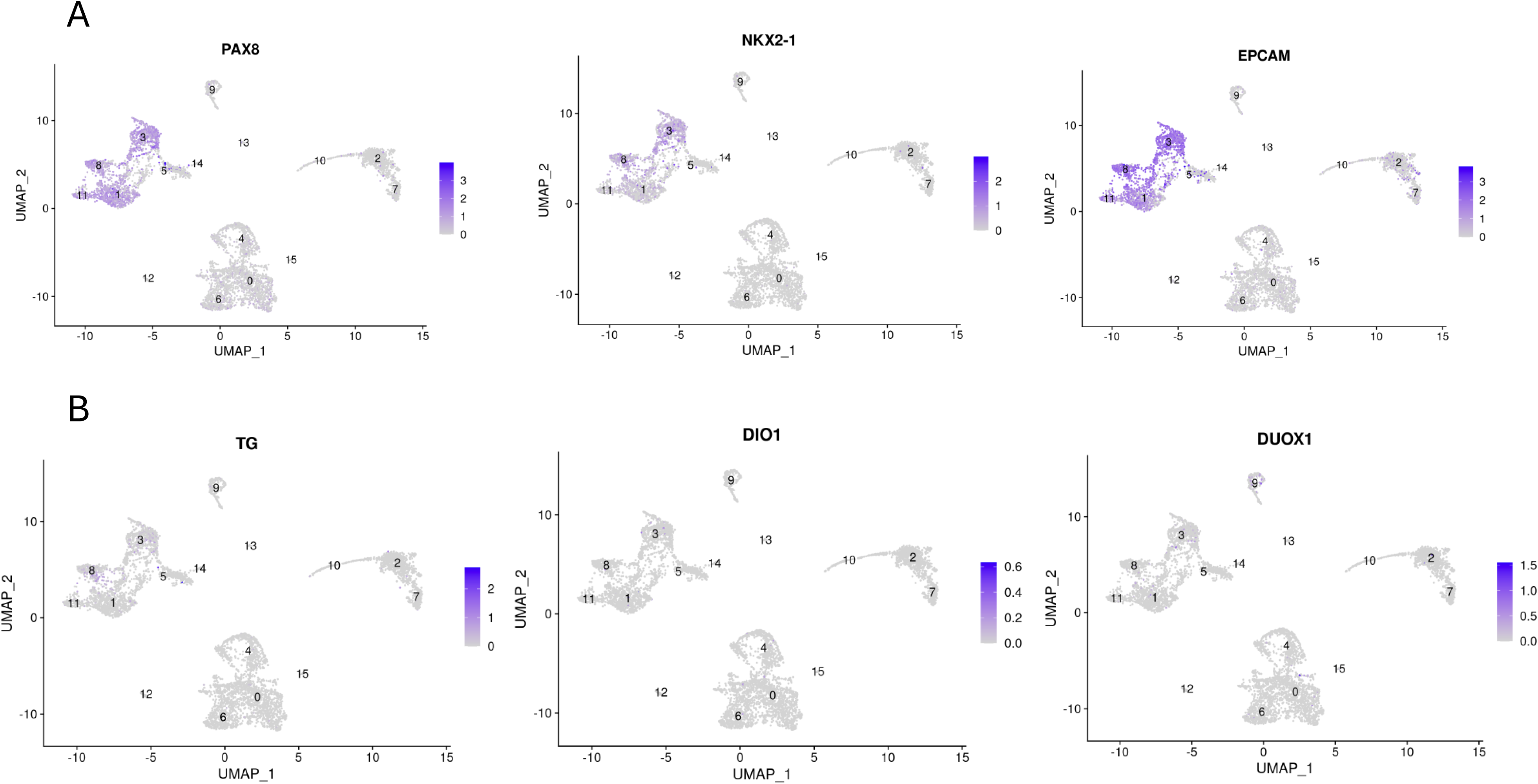
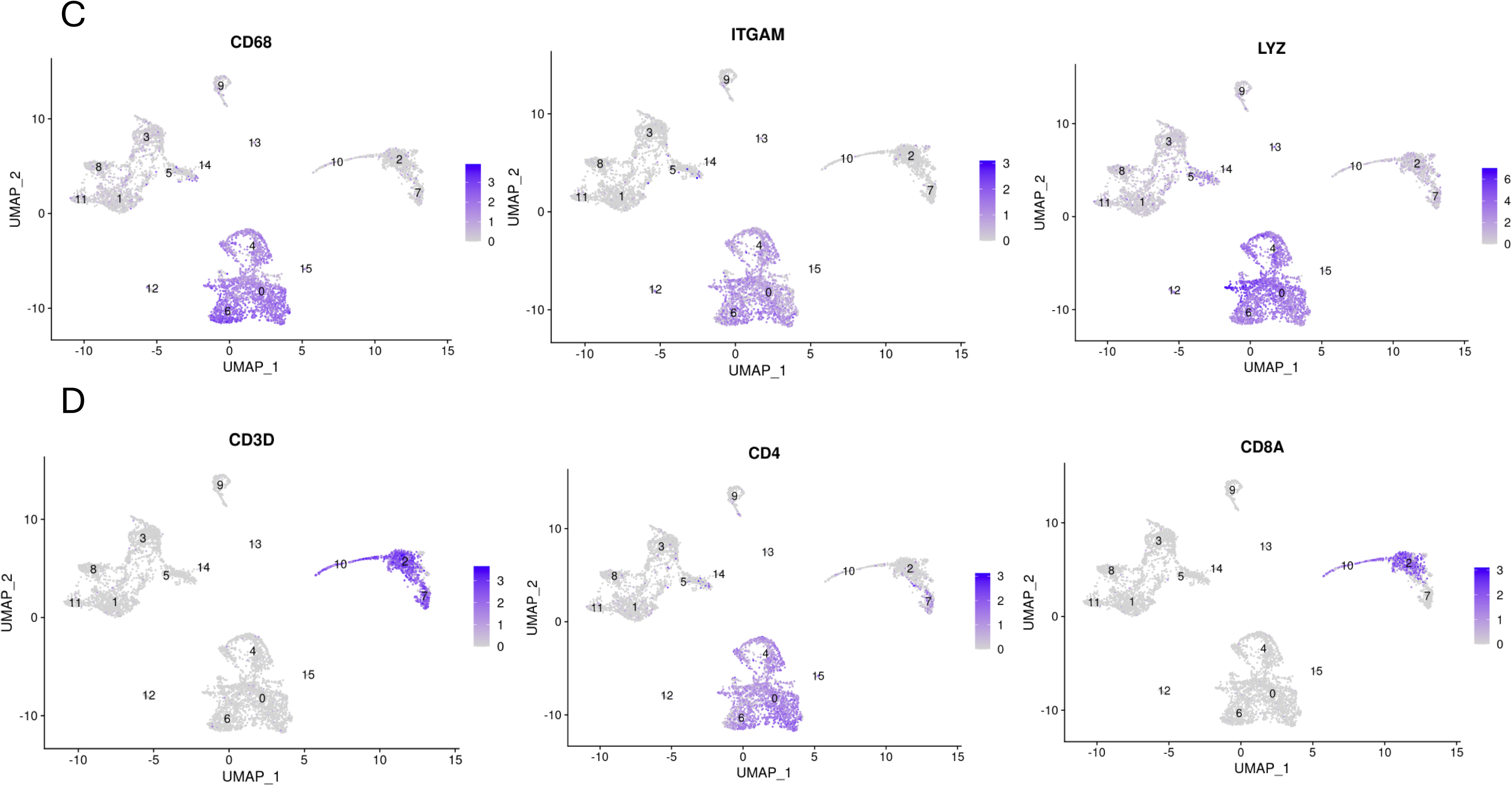
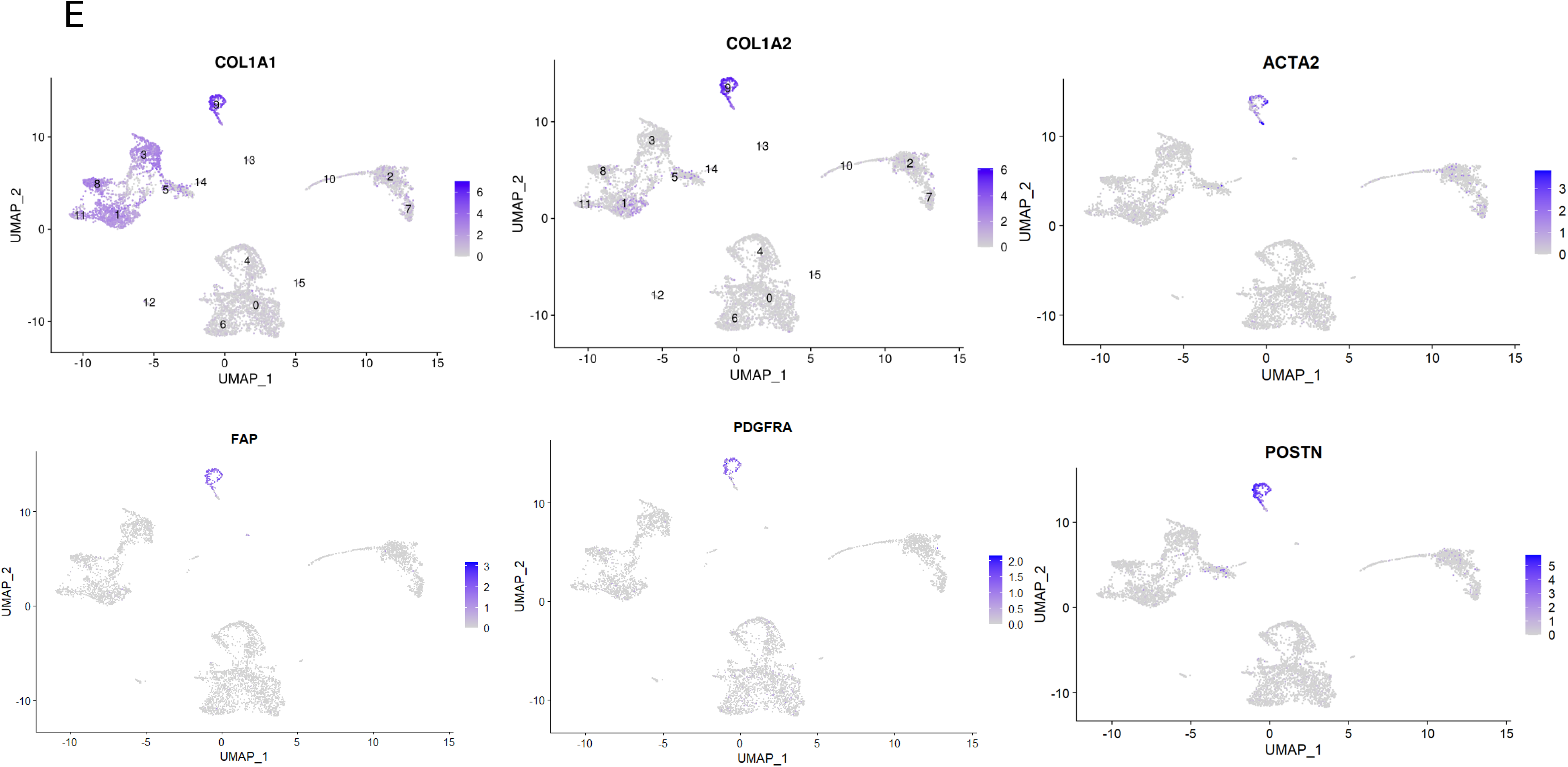
single cell RNA sequencing of a primary tumor from a patient with anaplastic thyroid cancer. A patient underwent resection of a primary ATC (*CCDC6-RET* fusion, c.1-124C>T *TERT* promoter mutation, *TP53* pR280K mutation). 5612 cells were sequenced, 27163 average number of UMI/cell and 4335 average number of genes per cell sequenced. mRNA markers were used to define cell types. A) markers of thyrocyte-derived and epithelial cells, B) thyrocyte differentiation markers, C) myeloid cell markers, D) T cell markers, E) CAF markers.

**Figure 2.**
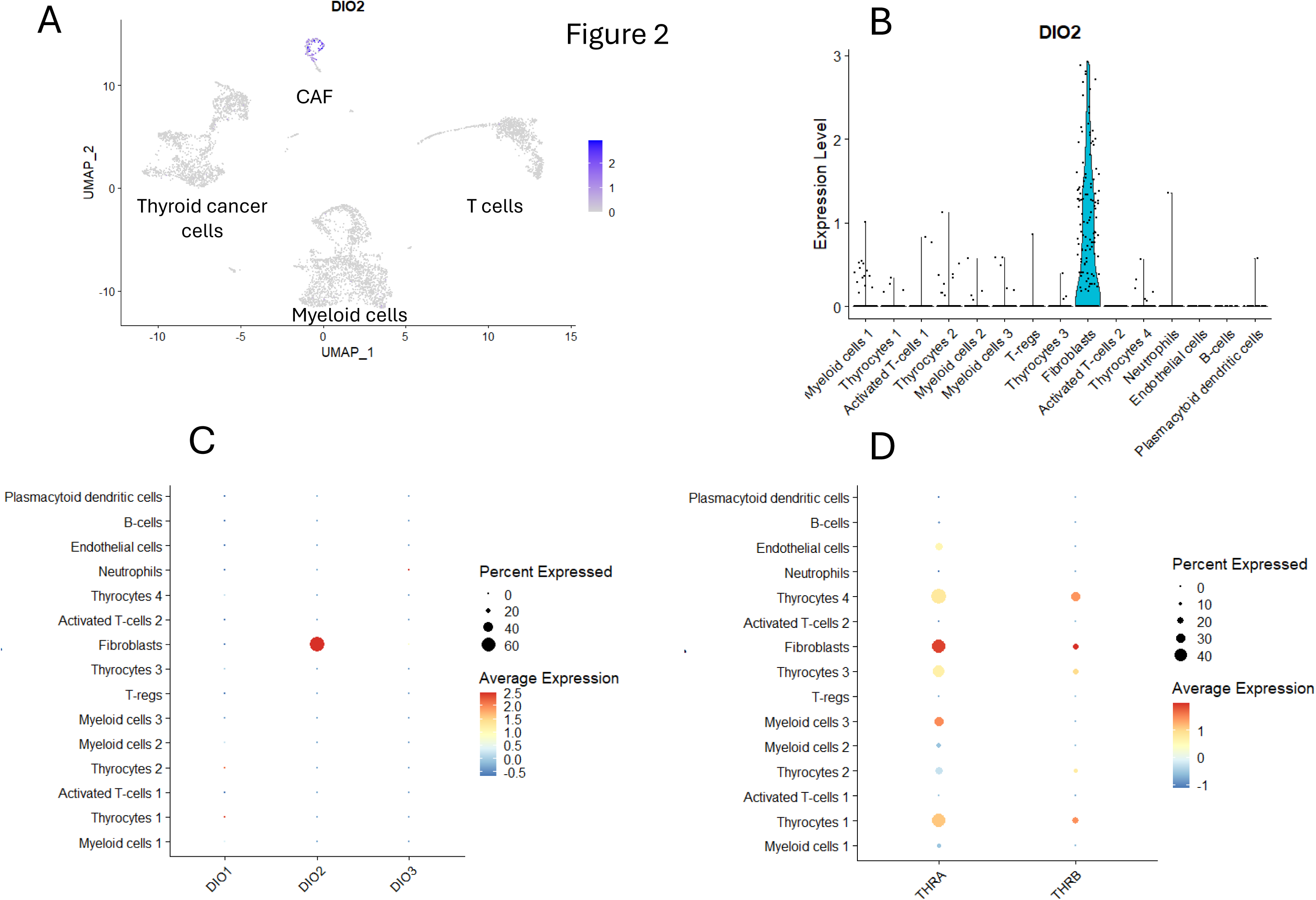
scRNAseq of ATC patient tumor reveals *DIO2* expression in CAFs. A) Feature plot (UMAP) shows expression of *DIO2* exclusively in the CAF cluster, B) Violin plots shows expression of *DIO2* exclusively in the CAF, C) Dot Plot comparing expression of *DIO1-3* in clusters. The size of the dot indicates percentage of cells expressing each mRNA and the color indicates the average level of expression. *DIO1* and *DIO3* are not expressed in any of the cell clusters, while *DIO2* is expressed exclusively in approximately 60% of the CAF, D) Dot Plot showing expression of the thyroid hormone receptors in cell clusters. *THRA* is expressed at the highest level in approximately 40% of the CAF and at lower levels in the thyroid cancer cells. *THRB* is expressed in a smaller percentage of CAF and thyroid cancer cells.

### DIO2 is expressed in CAF in a large scRNASeq thyroid cancer atlas

To determine if *DIO2* expression in CAF from our ATC patient is generalizable in advanced thyroid cancer, we examined expression of the deiodinases in stromal cells from a large scRNASeq Thyroid Atlas (21). This Atlas included 7 studies with 22 ATC samples, 39 PTC samples and 20 normal thyroid (paratumor) samples. The Atlas analysis included 423,733 cells and 24,463 stromal cells. These stromal cells included 5 major clusters: pericytes, inflammatory CAF (iCAF), myofibroblastic CAF (myCAF), vascular smooth muscle cells (vSMC) and APO-expressing perivascular-like cells (APOE+). Figure 3A shows UMAP and Violin plots of the stromal cells. *DIO2* is most highly expressed in a large number of myCAF cells, with lower expression in a smaller number of iCAF cells. The Dot plot in Figure 3B shows that *DIO2* is expressed in approximately 50% of myCAF and at lower levels in approximately 20% of iCAF. There is negligible expression of *DIO2* in the other stromal cell clusters. *DIO1* and *DIO3* have negligible expression in any of the stromal clusters. Figure 3C shows that *THRA* is expressed in a subpopulation of all of the stromal cells, while *THRB* is expressed at lower levels in fewer cells. These data confirm the results from our single patient ATC study and further define myCAF as the CAF subpopulation with the highest expression of *DIO2*.

**Figure 3.**
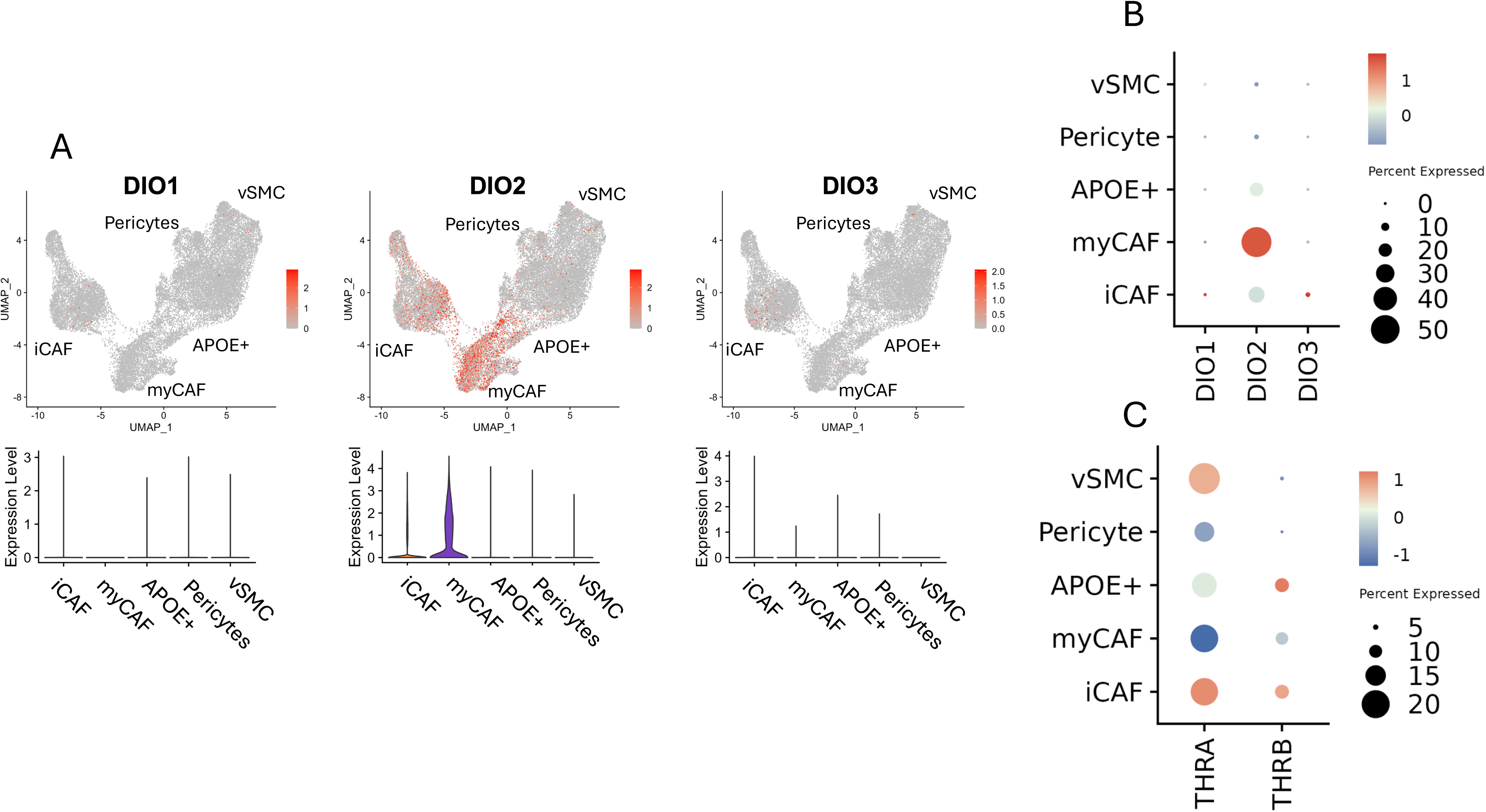
*DIO2* is expressed in CAF from a large scRNASeq thyroid cancer dataset. The scRNASeq Thyroid Atlas (21) is an integration of 81 thyroid tumor samples from 7 studies, which includes 39 PTC samples, 22 ATC samples and 20 normal thyroid/paratumor samples. The overall analysis is of 423,733 cells and the analysis for this figure was from 24,463 stromal cells. A) UMAP features plot (above) and Violin Plot (below) for *DIO1-3* of the thyroid atlas stromal cells. iCAF – inflammatory CAF, myCAF – myofibroblastic CAF, *APOE+* - APOE positive perivascular like cells, pericytes, vSMC – vascular smooth muscle cells. *DIO1* and *DIO3* are not expressed in these stromal cell clusters. *DIO2* is expressed primarily in the CAF clusters with highest expression in the myCAF cluster, B) Dot Plot of deiodinase expression in the different stromal cell clusters. *DIO2* is highly expressed in approximately 50% of myCAF, and expressed at lower levels in approximately 20% of myCAF.

We further subdivided the stromal cells by thyroid cancer type (ATC vs PTC) and normal thyroid tissue. Figure 4 shows that *DIO2* has the highest expression in the largest proportion of cells in myCAF from ATC samples, followed by myCAF from PTC samples. There are lower levels of *DIO2* expression in iCAF from PTC and ATC samples. As expected, there were very few myCAF in the normal (paratumor) tissues. These data suggest that DIO2 may play a functional role in myCAF from both ATC and PTC tumors.

**Figure 4.**
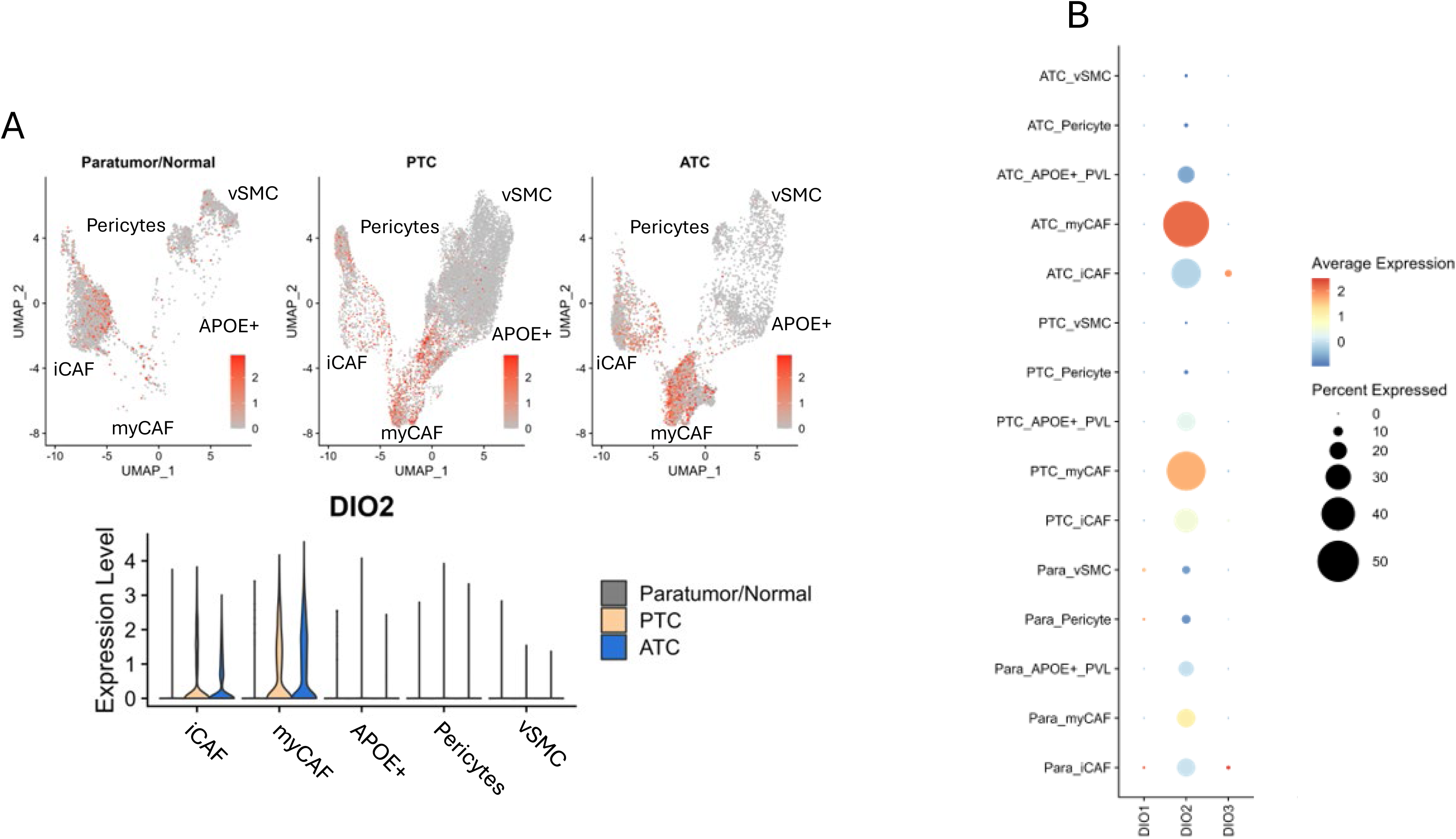
*DIO2* is most highly expressed in myCAF from ATC tumors but also expressed in myCAF from PTC tumors from a large scRNASeq thyroid cancer dataset. The scRNASeq Thyroid Atlas (21) is an integration of 81 thyroid tumor samples from 7 studies, which includes 39 PTC samples, 22 ATC samples and 20 normal thyroid/paratumor samples. The overall analysis is of 423,733 cells and the analysis for this figure was from 24,463 stromal cells subdivided into samples from normal thyroid tissues or paratumor tissue (‘normal’ thyroid near the tumor), PTC and ATC. A) UMAP features plot (above) and Violin Plot (below) for *DIO1-3* of the thyroid atlas stromal cells. iCAF – inflammatory CAF, myCAF – myofibroblastic CAF, *APOE+* - APOE positive perivascular like cells, pericytes, vSMC – vascular smooth muscle cells. Paratumor/normal thyroid tissue has fewer pericytes and *APOE+* cells that PTC or ATC and almost no myCAF. ATC has the highest amount of myCAF. *DIO2* is expressed in myCAF from ATC and PTC and at lower levels in iCAF from ATC and PTC, B) Dot Plot of *DIO1-3* expression in the different clusters from the three types of tissue. *DIO1* is not expressed in these clusters. *DIO3* may be expressed in a very small fraction of ATC and normal thyroid iCAF. *DIO2* is most highly expressed in approximately 50% of ATC myCAF and is expressed in approximately 40% of PTC myCAF. *DIO2* is expressed at low levels in approximately 30% of ATC iCAF and 20% of PTC iCAF. Expression is lower in other clusters.

### DIO2 is expressed in CAF from different cancer types

To test the hypothesis that DIO2 is expressed in CAF from many different solid tumors, we performed scRNASeq analyses on publicly available datasets from six different tumor types including breast carcinoma, head and neck squamous cell carcinoma (HNSCC), lung adenocarcinoma, colorectal carcinoma, prostate carcinoma and pancreatic adenocarcinoma (13–18). *DIO2* was exclusively expressed in the CAF from these different tumors. The Dot Plot in Figure 5 shows that *DIO2* is expressed in 8-40% of CAF from these different tumors, suggesting that DIO2 and local production of T3 may play a role in CAF function across multiple tumor types. As was seen in thyroid cancer, *DIO1* and *DIO3* have negligible expression in the different solid tumors, although 10% of CAF from PDAC appear to express some *DIO3*.

**Figure 5.**
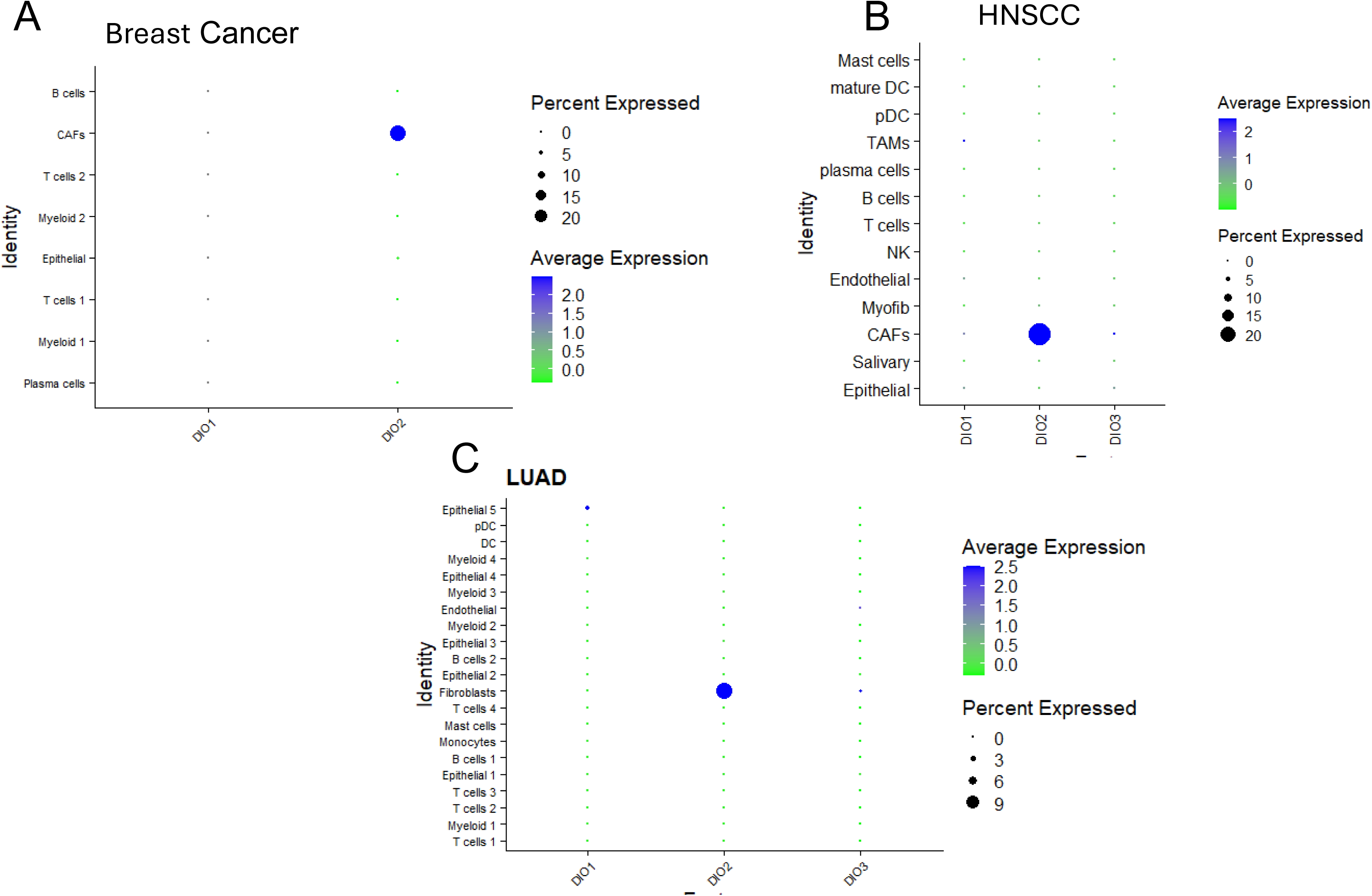
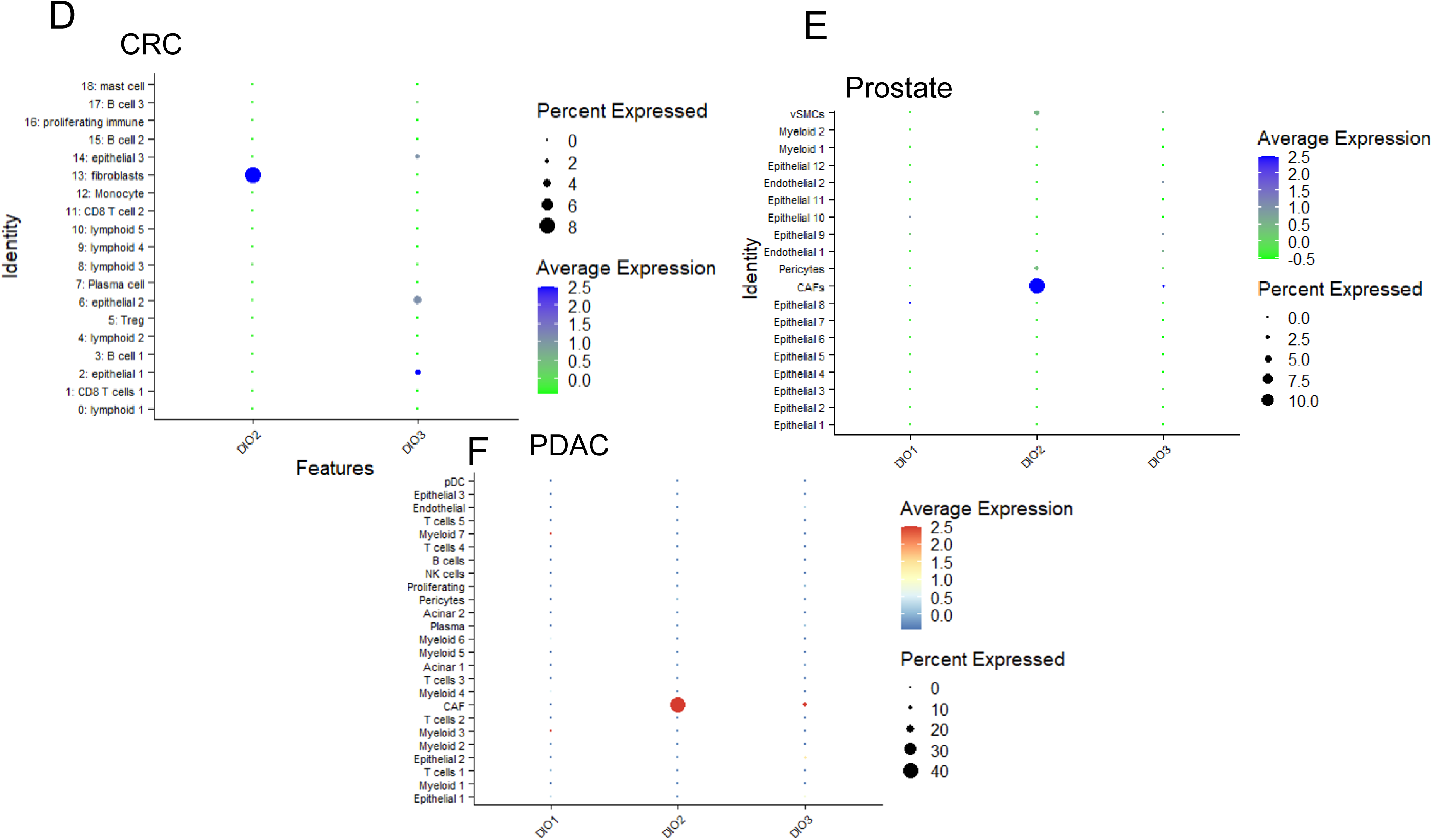
CAF-specific *DIO2* expression is identified in many cancer types. scRNASeq data was extracted and analyzed from different cancer types. Dot plots show average expression of *DIO1, 2, and 3* in A) breast cancer, B) head and neck squamous cell cancer (HNSCC), C) lung adenocarcinoma (LUAD), D) colorectal carcinoma (CRC), E) prostate cancer F) pancreatic ductal adenocarcinoma (PDAC) from scRNAseq datasets. Color indicates average gene expression level and dot size represents the percent of deiodinase expressing cells in each cluster. *DIO3* and *DIO1* expression are not shown in breast and colorectal cancers, respectively, due to a lack of any detectable expression in the tumor samples. *DIO1 and 3* are not expressed or expressed at very low levels in the different cancers. *DIO3* is expressed in approximately 10% of PDAC CAF. *DIO2* is expressed in 40% of CAF from PDAC, 20% of CAF from breast cancer and HNSCC, 10% of CAF from prostate cancer, 9% of CAF from LUAD, and 8% of CAF from CRC.

### *DIO2 expression correlates with myCAF markers in large bulk RNASeq datasets from* different cancers

To further study the relationship between CAF and DIO2 expression in large tumor datasets, we used the ORIEN database which has clinical data, RNA sequencing and whole exome sequencing on many tumor types from >25,000 patients. To determine the portion of myCAF and iCAF in different tumors, we first compared the expression of six myCAF markers (*FAP, POSTN, COL1A2, ACTA2, PDGFRA, MMP11*) in different cancer type datasets with >500 tumors. The overall best correlation between two myCAF markers was FAP and COL1A2 among the different tumor types. Figure 6 shows six tumor types with strong correlation between *FAP* and *COL1A2* expression. There are specific tumors (each dot) with relatively high proportions of myCAF (upper right of each graph) and tumors with relatively low proportions of myCAF (lower left). Figure 7 shows correlation of between *FAP* (myCAF) and *DIO2* and *DIO3* expression in the six tumor types with the best correlation between *FAP* and *DIO2* expression.

**Figure 6.**
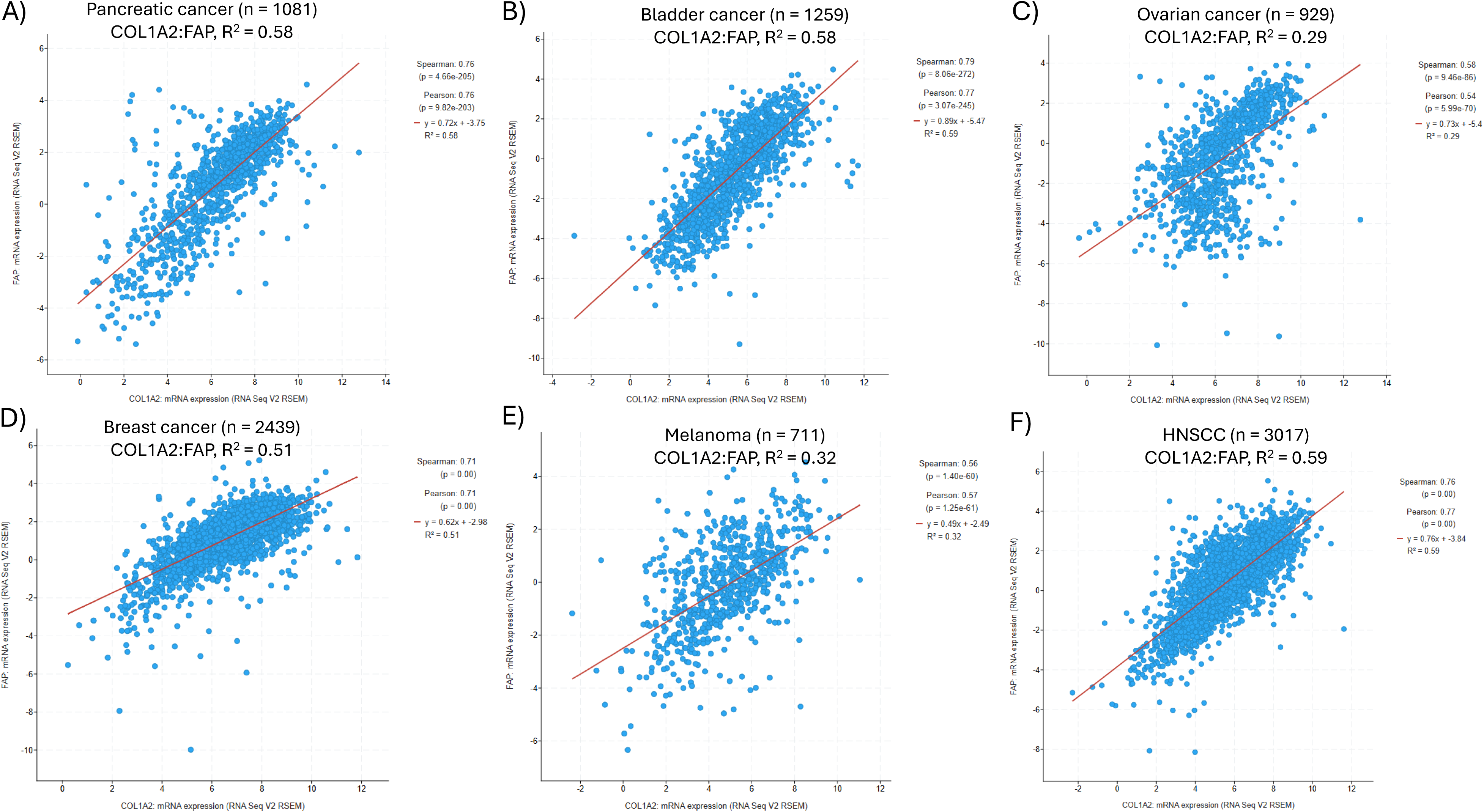
The myCAF markers *FAP* and *COL1A2* show a strong correlation in bulk RNASeq from six different tumor types. The Oncology Research Information Exchange Network (ORIEN) database was queried for mRNA expression across multiple tumor types. Two myCAF markers, *FAP* and *COL1A2*, were compared as an estimate of proportion of myCAFs in each tumor (represented by dots). A) Pancreatic cancer (n=1081), B) Bladder cancer (n=1259), C) Ovarian cancer (n=929), D) Breast cancer (n=2439), E) Melanoma (n=711), F) HNSCC (n=3017). Most tumor types showed a strong correlation (R^2^ > 0.50) between the two myCAF markers.

**Figure 7.**
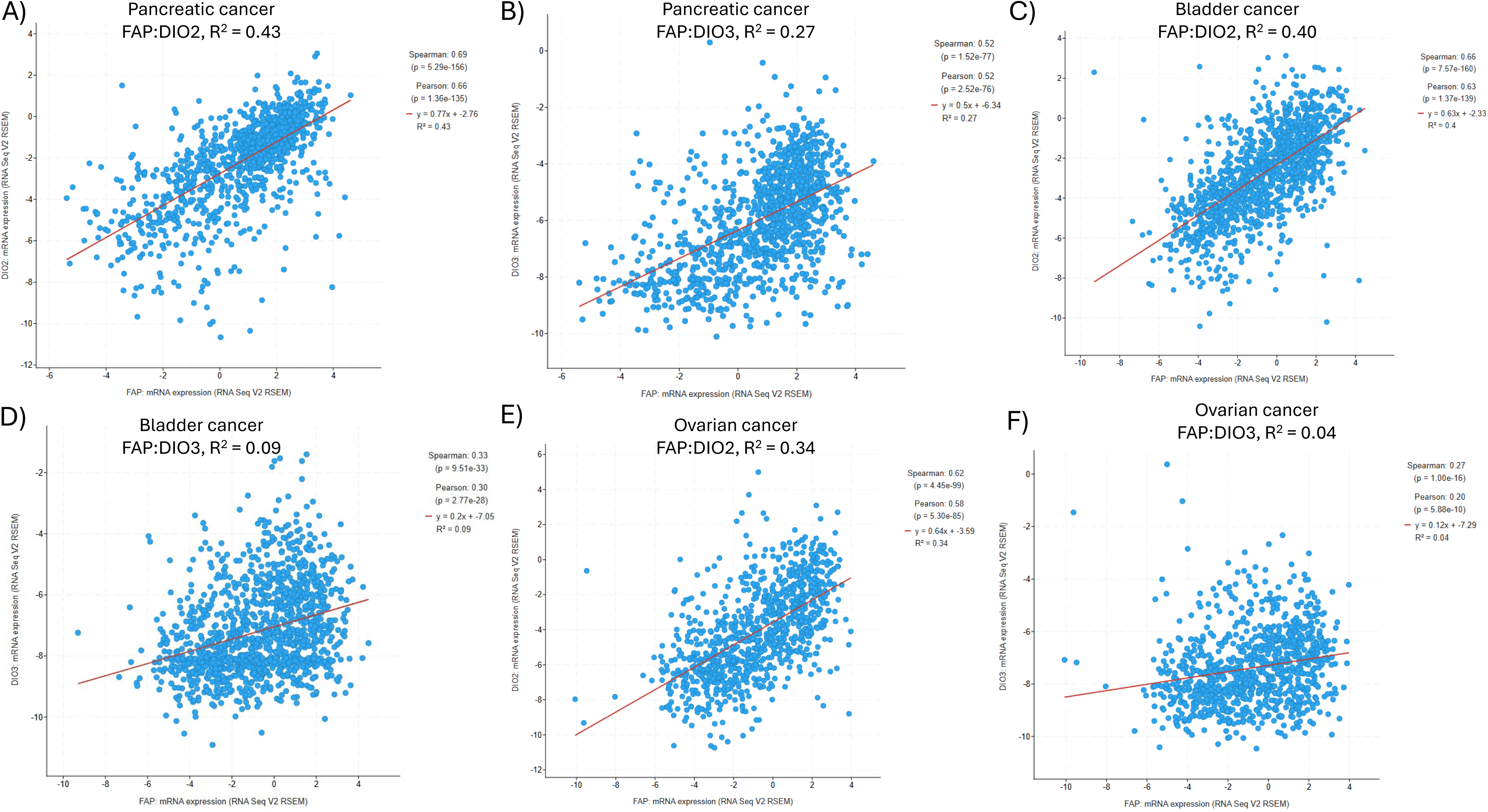
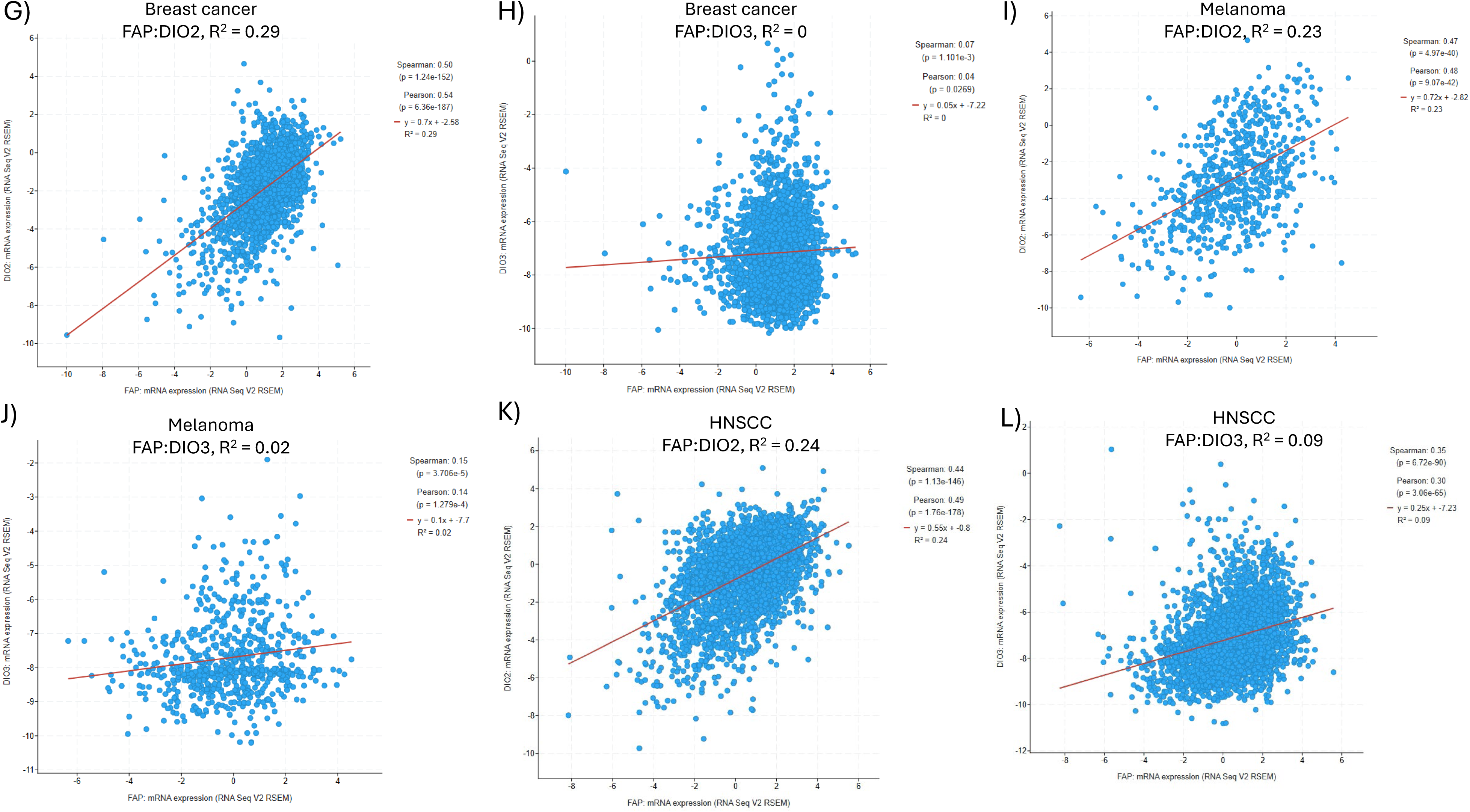
The myCAF marker *FAP* has a stronger correlation with DIO2 expression compared with *DIO3* expression. The Oncology Research Information Exchange Network (ORIEN) database was queried for mRNA expression across multiple tumor types. The myCAF marker *FAP* expression was correlated with expression of *DIO2* in each tumor (represented by dots). There was negligible expression of *DIO1* in these tumors (data not shown). A-B) Pancreatic cancer (n=1081), C-D) Bladder cancer (n=1259), E-F) Ovarian cancer (n=929), G-H) Breast cancer (n=2439), I-J) Melanoma (n=711), K-L) HNSCC (n=3017).

Pancreatic cancer had the strongest correlation between *FAP* and *DIO2* expression (Fig 7A, R^2^ = 0.43), and less of a correlation between *FAP* and *DIO3* expression (Fig 7B, R^2^ = 0.27), although still significant. *DIO2* mRNA levels were much higher in all tumors compared with *DIO3* levels. There was no correlation between FAP and *DIO1* expression in any of the tumor types (R^2^ ≤ 0.04, data not shown). The other five tumor types (bladder, ovarian, breast, melanoma and HNSCC) showed good correlation between *FAP* and *DIO2* expression, and much lower correlation between *FAP* and *DIO3* expression. We also observed strong correlation of expression between *COL1A2* or *POSTN* and *DIO2* in these tumors, suggesting that myCAF express *DIO2* in many tumor types.

We performed the same analyses using iCAF markers (*CXCL12, DPT, APOD*) and found much lower correlations between these CAF markers and *DIO2* expression (R^2^ = 0.01-0.20, data not shown) and *DIO3* expression (R^2^ = 0.01-0.20, data not shown). These bulk mRNASeq data from large numbers of tumors support the scRNASeq data in from a smaller number of cancers showing higher *DIO2* expression in myCAF compared with iCAF and higher expression of *DIO2* in CAF compared with *DIO3*.

Thyroid cancer showed an inverse correlation between myCAF markers and *DIO2* expression (Fig 8). This is likely due to the expression of DIO2 in normal thyrocytes and differentiated thyroid cancer, with loss of expression in more dedifferentiated tumors that are associated with higher myCAF in the tumors. Using different datasets, we show that *DIO2* has highest expression in normal thyroid tissue, with lower *DIO2* expression as the tumors become dedifferentiated into ATC (Fig 9). Interestingly, ATC cells lines have much lower *DIO2* expression than ATC tumors, likely due to the absence of CAF in ATC cells lines.

**Figure 8.**
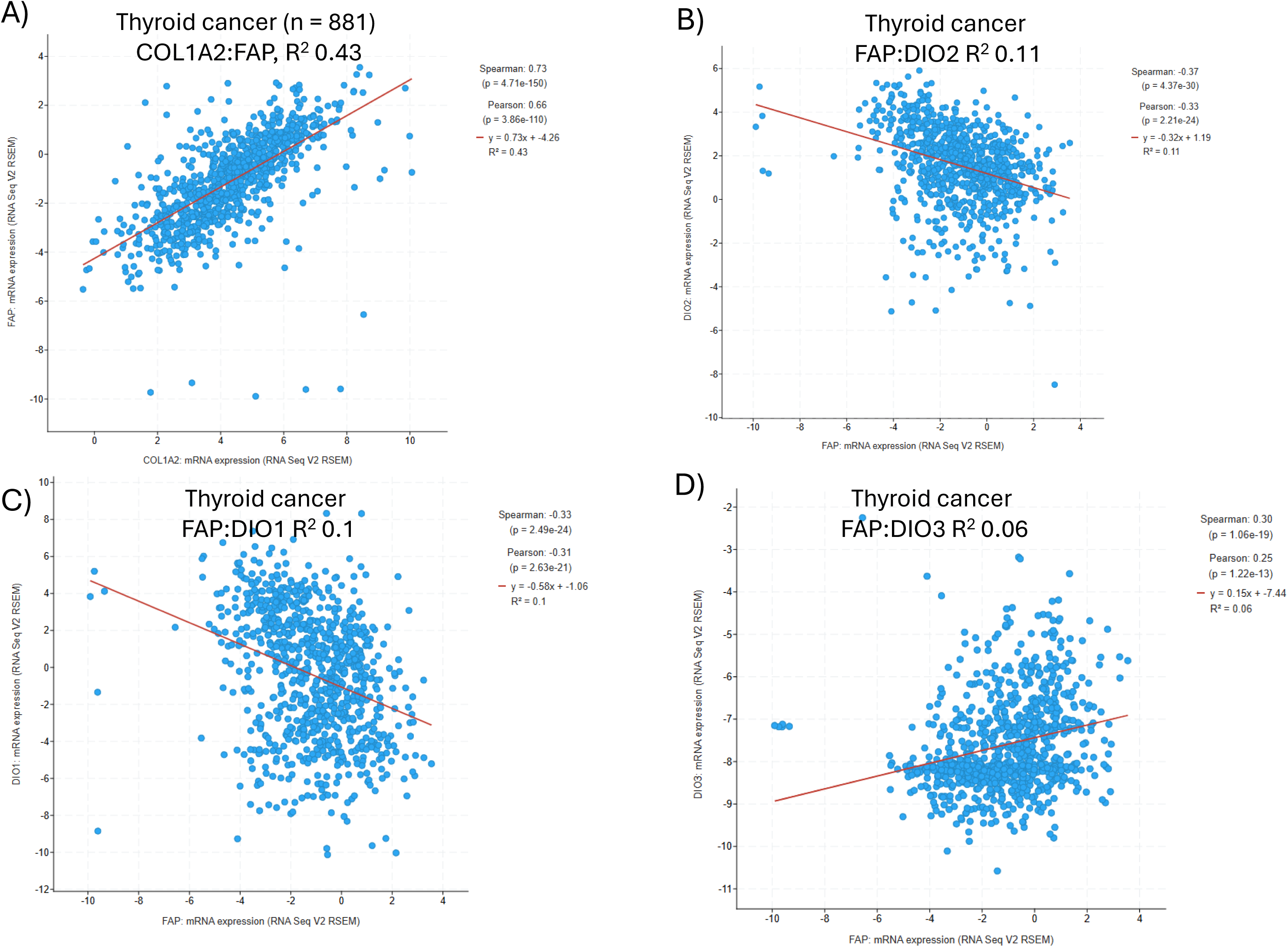
The myCAF markers *FAP* and *COL1A2* show a good correlation in thyroid cancer tumors and *FAP* shows a weak inverse correlation with *DIO2* and *DIO1* in these tumors. The Oncology Research Information Exchange Network (ORIEN) database was queried for mRNA expression in thyroid cancers (n=881). The myCAF marker *FAP* expression was correlated with expression of A) *COL1A2*, B) *DIO2*, C) *DIO1*, and D) *DIO3* in each tumor (represented by dots).

**Figure 9.**
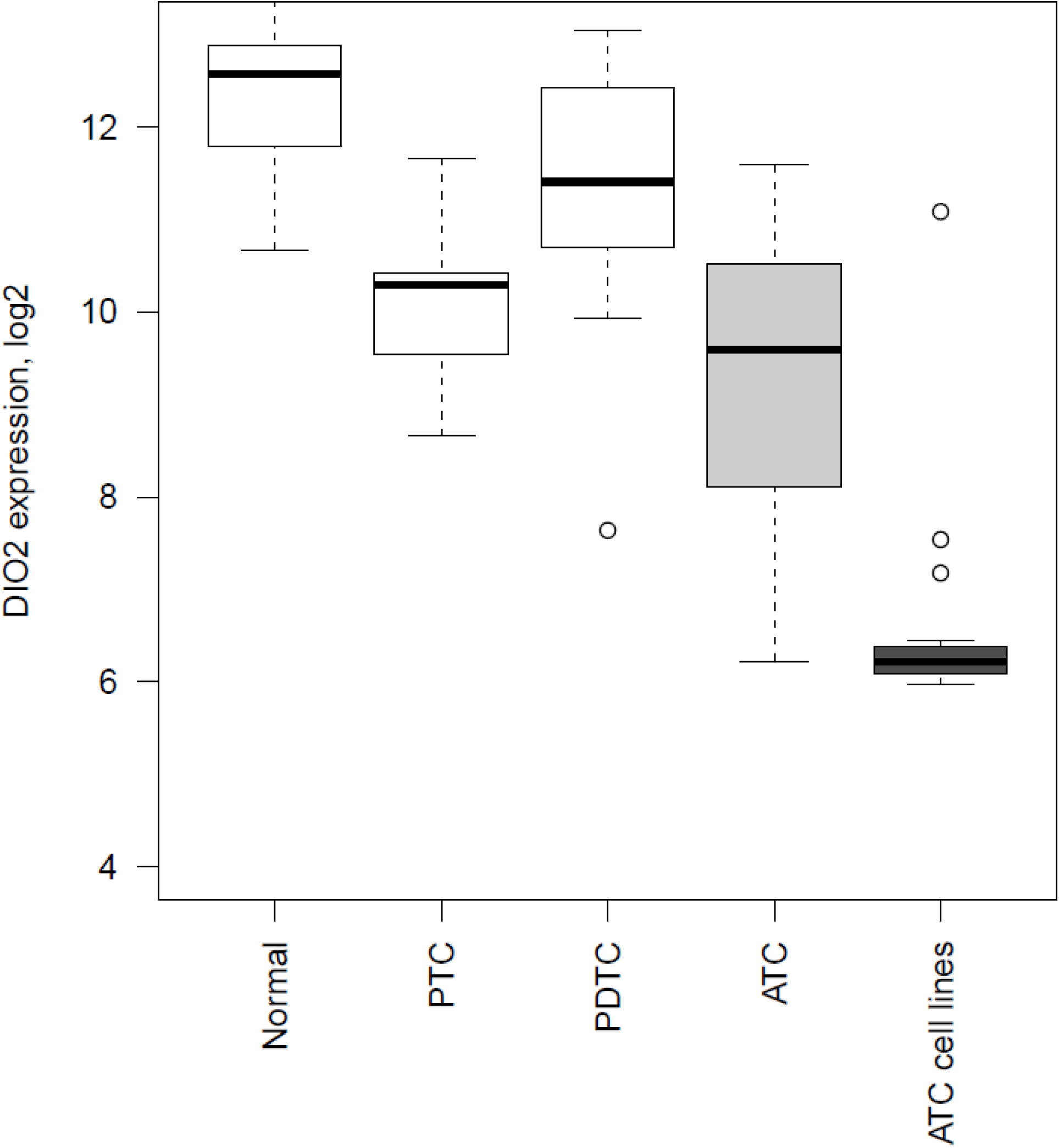
*DIO2* mRNA is variably expressed across different thyroid tissue types. Raw gene expression data (Affymetrix GeneChip Human Genome U133 Plus 2.0) for normal thyroid tissue, papillary thyroid cancer (PTC), poorly differentiated thyroid cancer (PDTC), and anaplastic thyroid cancer (ATC) were downloaded from Gene Expression Omnibus (22) and GSM2024835 (23). Thyroid cancer cell line gene expression data was acquired from our prior publication (24). To reduce batch effect, background subtraction, normalization and summarizing probe sets were done jointly for CEL files from all three sources.

### *myCAF marker and DIO2 expression is associated with overall survival in large bulk RNASeq* datasets from different cancers

To determine whether myCAF markers and *DIO2* mRNA expression were associated with overall survival (OS) across different cancer types, we used the ORIEN database. We first generated Kaplan-Meier plots with a binary cutoff for the myCAF marker, *FAP*, using median mRNA expression. High *FAP* expression was significantly associated with worse OS for patients with pancreatic, bladder, ovarian, thyroid cancers and HNSCC, suggesting that higher amounts of myCAF in these cancers are associated with worse OS (Fig 10). Interestingly, the opposite association was seen in breast cancer and melanoma (Fig 10D and data not shown).

**Figure 10.**
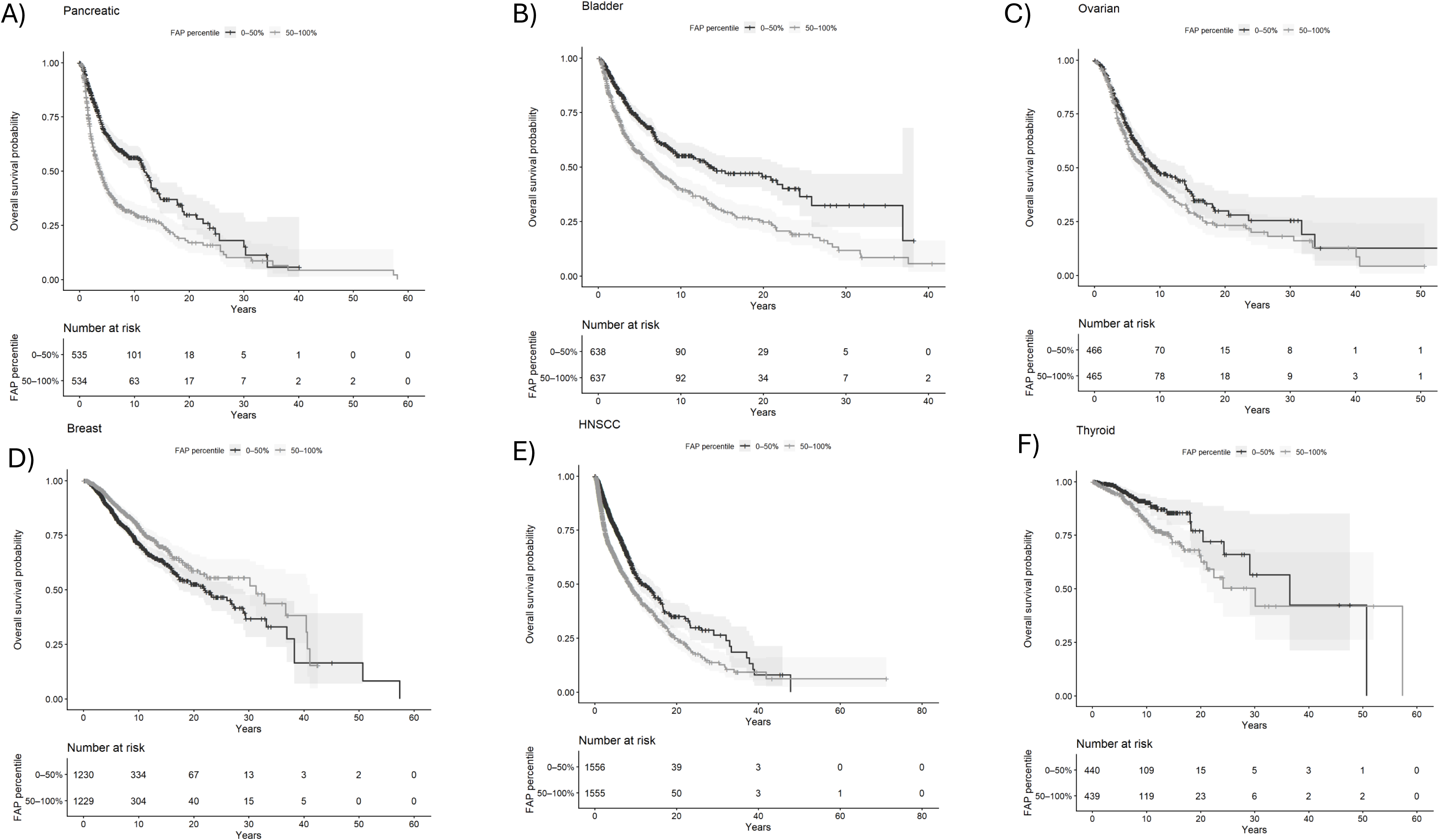
**Expression of the myCAF marker, *FAP*, is associated with worse overall survival across many cancer types**. Kaplan-Meier plots were generated for the association of high or low levels of *FAP* RNA with overall survival (OS) across multiple cancer types in the ORIEN database. High versus low levels of *FAP* were divided by the median expression levels in each tumor type. A) Pancreatic cancer (n=1069), B) Bladder cancer (n=1275), C) Ovarian cancer (n=931), D) Breast cancer (n=2459), E) HNSCC (n=3111), F) Thyroid cancer (n=879). *FAP* RNA levels were associated with worse OS in all tumor types except breast cancer (inverse association) and melanoma (inverse association, data not shown).

*DIO2* expression was significantly associated with worse OS in patients with pancreatic and bladder cancers (Fig 11A and B). These two tumor types showed the best correlation between the myCAF marker *FAP* and *DIO2* expression (Fig 7). *DIO2* expression was not associated with OS in ovarian and breast cancers or melanoma and HNSCC (Fig 11 C-E and data not shown). Interestingly, *DIO2* expression was inversely associated with OS in thyroid cancer (Fig 11F). Thyroid and breast tissue express *DIO2* and these cancers have reduced levels of *DIO2* as the epithelial cells dedifferentiate (Fig 9, thyroid).

**Figure 11.**
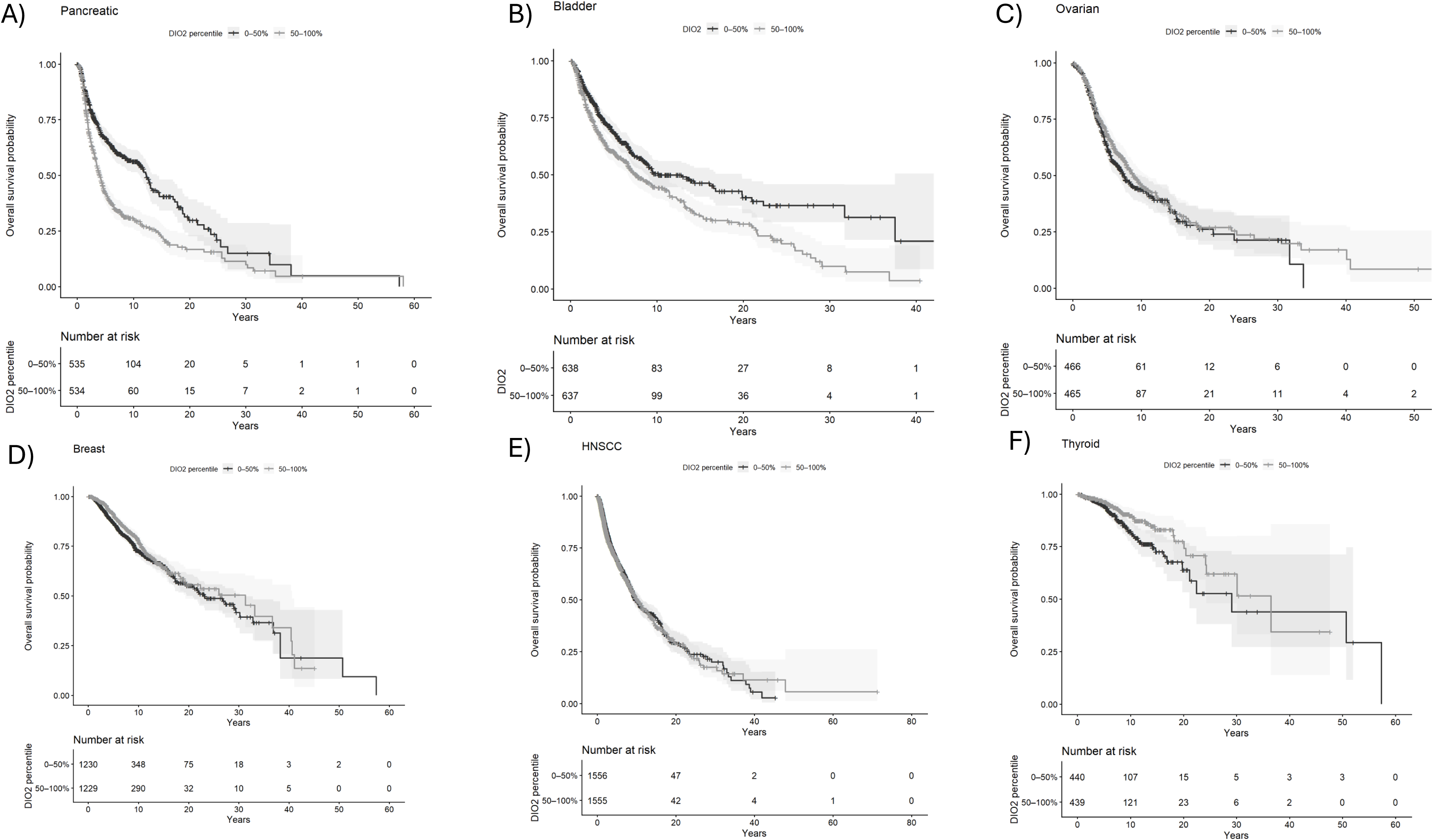
Expression of *DIO2* is associated with worse overall survival in some cancer types. Kaplan-Meier plots were generated for the association of high or low levels of *DIO2* RNA with overall survival (OS) across multiple cancer types in the ORIEN database. High versus low levels of *DIO2* were divided by the median expression levels in each tumor type. A) pancreatic cancer (n=1069), B) bladder cancer (n=1275), C) ovarian cancer (n=931), D) breast cancer (n=2459), E) HNSCC (n=3111), F) thyroid cancer (n=879). *DIO2* RNA levels were associated with worse OS in pancreatic and bladder cancers, but not in ovarian and breast cancers, melanoma or HNSCC. OS was inversely correlated with *DIO2* expression in thyroid cancer.

As a complementary analysis, we performed Cox proportional hazards modeling using each CAF marker and *DIO2* as a continuous variable. The hazard ratio (HR) was the incremental change is OS for a one-unit change in mRNA level for each marker. Higher *FAP* expression was significantly associated with worse OS for patients with pancreatic, bladder, ovarian, thyroid cancers and HNSCC, consistent with the Kaplan-Meier analysis suggesting that higher amounts of myCAF in these cancers are associated with worse OS (Fig 12A). The opposite association was seen for breast cancer and there was a nonsignificant trend for melanoma. We tested a second myCAF marker, POSTN, and found similar results except for no associations in melanoma and ovarian cancer (Fig 12B). To determine if the association was also seen with an iCAF marker, we repeated the analysis using the iCAF marker *CXCL12* (Fig 12C). There was a significant association seen for ovarian cancer, with a similar HR to the myCAF markers. While a significant association held for pancreatic cancer, the HR was smaller than for myCAF markers. Interestingly, there was an inverse association for three of the cancers (breast, HNSCC, melanoma), while there was no association seen for bladder and thyroid cancers. It appears that the myCAF markers are more strongly associated with OS in most of the cancers examined. *DIO2* expression was significantly associated with worse OS in pancreatic and bladder cancers, and inversely associated with breast and thyroid cancers, likely due to the expression of DIO2 in normal breast and thyroid tissue (Fig 12D). There was no association of *DIO2* expression and OS for HNSCC, melanoma or ovarian cancer.

**Figure 12.**
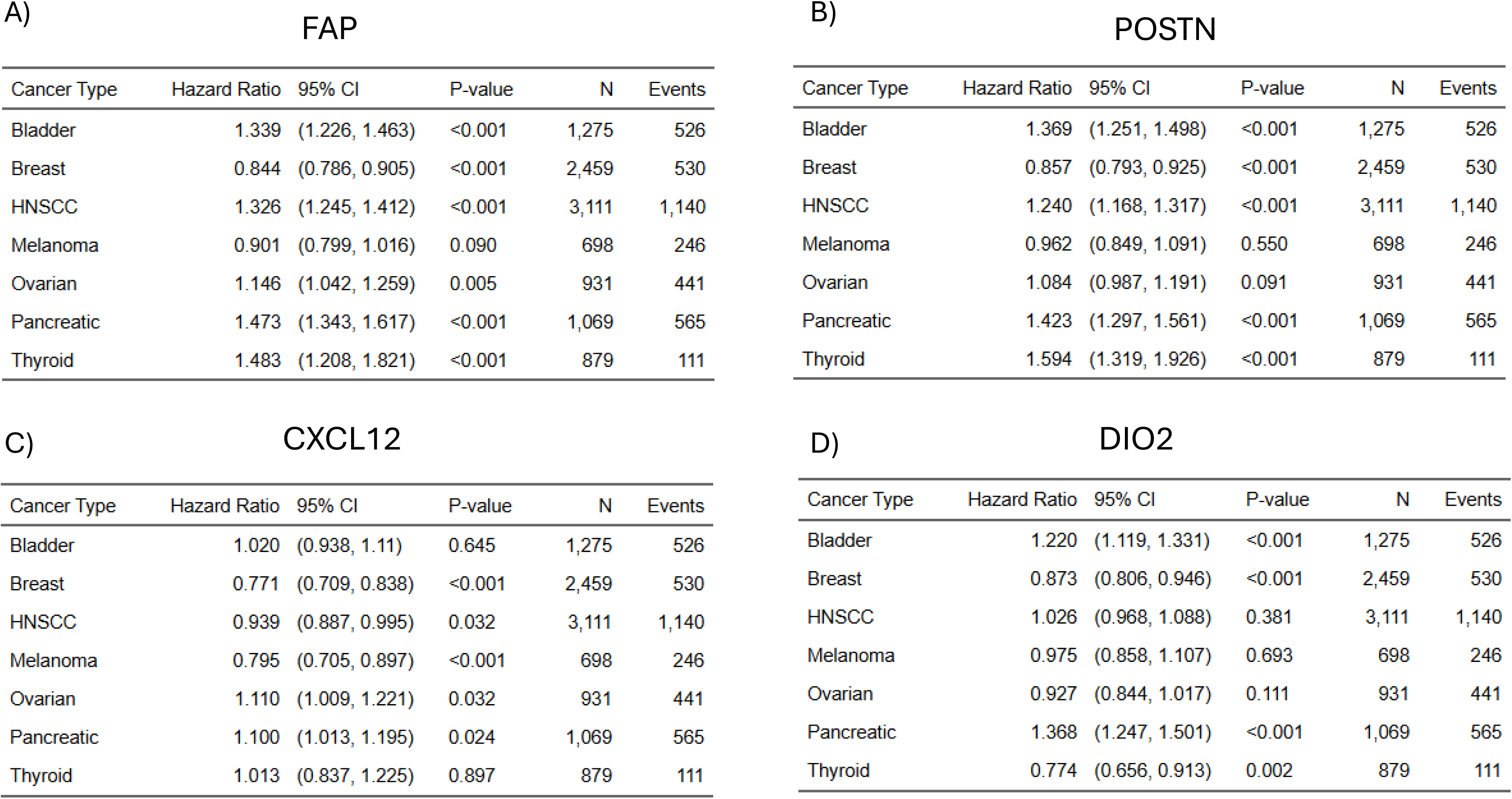
Cox proportional hazards analysis comparing the association of overall survival (OS) with different biomarkers across different types of cancer. Cox proportional hazards (CoxPH) modeling of OS was conducted using each CAF marker and DIO2 as a continuous variable for different cancers. Hazard ratios reflect a one-SD increase in biomarker expression. An HR greater than 1 indicates that the hazard of death increases as the biomarker increases, whereas an HR less than 1 indicates that the hazard of death decreases as the biomarker increases. Ninety-five percent confidence intervals (95% CI) around the HR and corresponding p-values are also presented. N denotes the number of subjects included in each model, and Events denotes the number of observed deaths used to estimate the model. Please note that the SD is calculated separately within each cancer type and biomarker. A) *FAP*, B) *POSTN*, C), *CXCL12*, D) *DIO2*.

### The TBP mouse is a good model to study Dio2 function in CAF

To study the effects of Dio2 in CAF, we utilized the TBP (***T****PO-CreERT2/LSL- **B**rafV600E/wt/Tr**p**53*^Δ*ex2-10/*Δ*ex2-10*^) mouse model. After induction with tamoxifen, these mice form thyroid tumors with components of PTC in smaller tumors and ATC in larger tumors over 14-30 weeks (Fig 10A). These tumors are highly fibrotic as noted with Col1a1 staining in Figures 10B and C. We performed scRNASeq on an ATC tumor from a female mouse harvested 20 weeks after tamoxifen induction. UMAP plot (Fig 10D) shows that the tumors are predominantly composed of thyroid tumor cells, myeloid cells and fibroblasts, with smaller components of T cells, neutrophils and endothelial cells. As seen in the human tumors, *Dio2* expression is restricted to the CAF (Fig 10D and E). *Dio1*is exclusively expressed in the differentiated thyrocytes, as expected. Fig 10F shows that the thyroid hormone receptor, *Tra*, has highest expression in the CAF, suggesting that T4 to T3 conversion in CAF will have significant thyroid hormone action effects directly in the CAF. The other thyroid hormone receptor, *Trb*, has very little expression in this tumor. The antibiotic, cefuroxime, has been shown to bind to Dio2 and reduce D2 activity in mouse tissues (27). We treated TBP mice with daily IP injection of 900 mg/kg/day cefuroxime or vehicle for 15 days. Cefuroxime reduced D2 activity by approximately 50% in mouse thyroid tumors (Fig 10G). There have been a few reports that multikinase inhibitors (MKI, selpercatinib and vandetinib) inhibited D2 activity in MSTO cells (fibroblast-like) and skeletal muscle fibroadipogenic progenitor (FAP) cells (30, 31). Interestingly, other MKI (pralsetinib and cabozantinib) did not inhibit D2 activity, suggesting that a specific signaling pathway is inhibited. The MKI lenvatinib is a potent inhibitor of tumor growth in patients with radioiodine refractory DTC (32). This MKI has not been tested for inhibition of D2 activity. Figure 10G shows that lenvatinib inhibits D2 activity in TBP mouse tumors in a dose-dependent manner. We have previously shown that lenvatinib reduces tumor growth in these mice (26). Inhibition of D2 activity may play a mechanistic role in the anti-tumor activity of lenvatinib.

A previous study showed that *Dio2* can be expressed in different cell types in the TME, especially when *Dio2* is lost in the fibroblasts (10). To determine the effect of complete Dio2 suppression in the TME and mimic robust pharmacologic suppression of D2 activity in the animal, we generated *TBP-Dio2 WT* and *TBP-Dio2 KO* littermates. *Dio2* KO mice continue to generate T3 through *Dio1* (28). Mice were given 0.2 mg/L LT4 in the drinking water since TBP mice have poor thyroid function and become hypothyroid. Figure 14 shows that *TBP-Dio2* KO mice have significantly slower thyroid tumor growth compared to *TBP-Dio2* WT littermates (p = 0.0022), making this an excellent *in vivo* model to study the effects of Dio2 in the TME.

**Figure 13.**
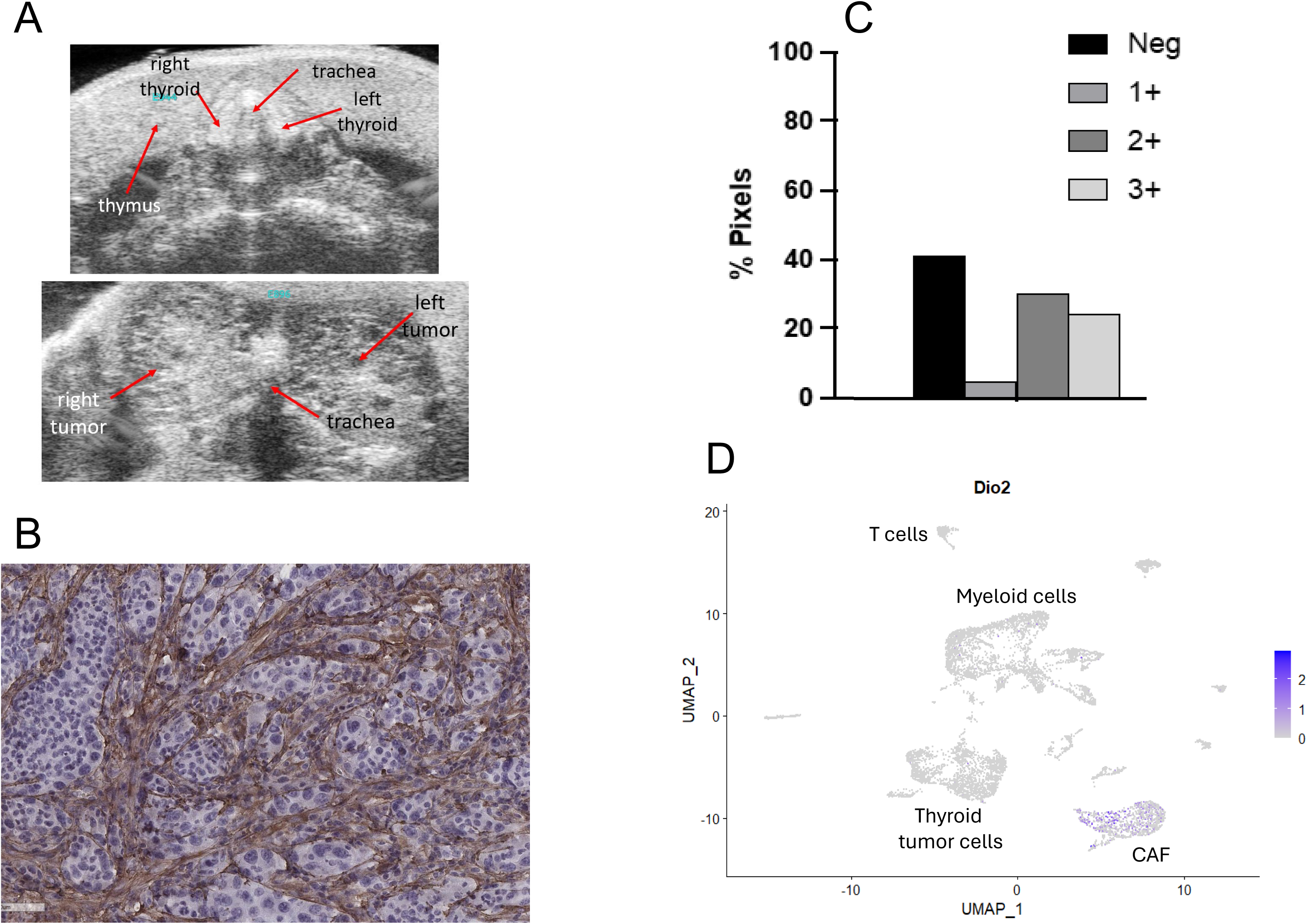
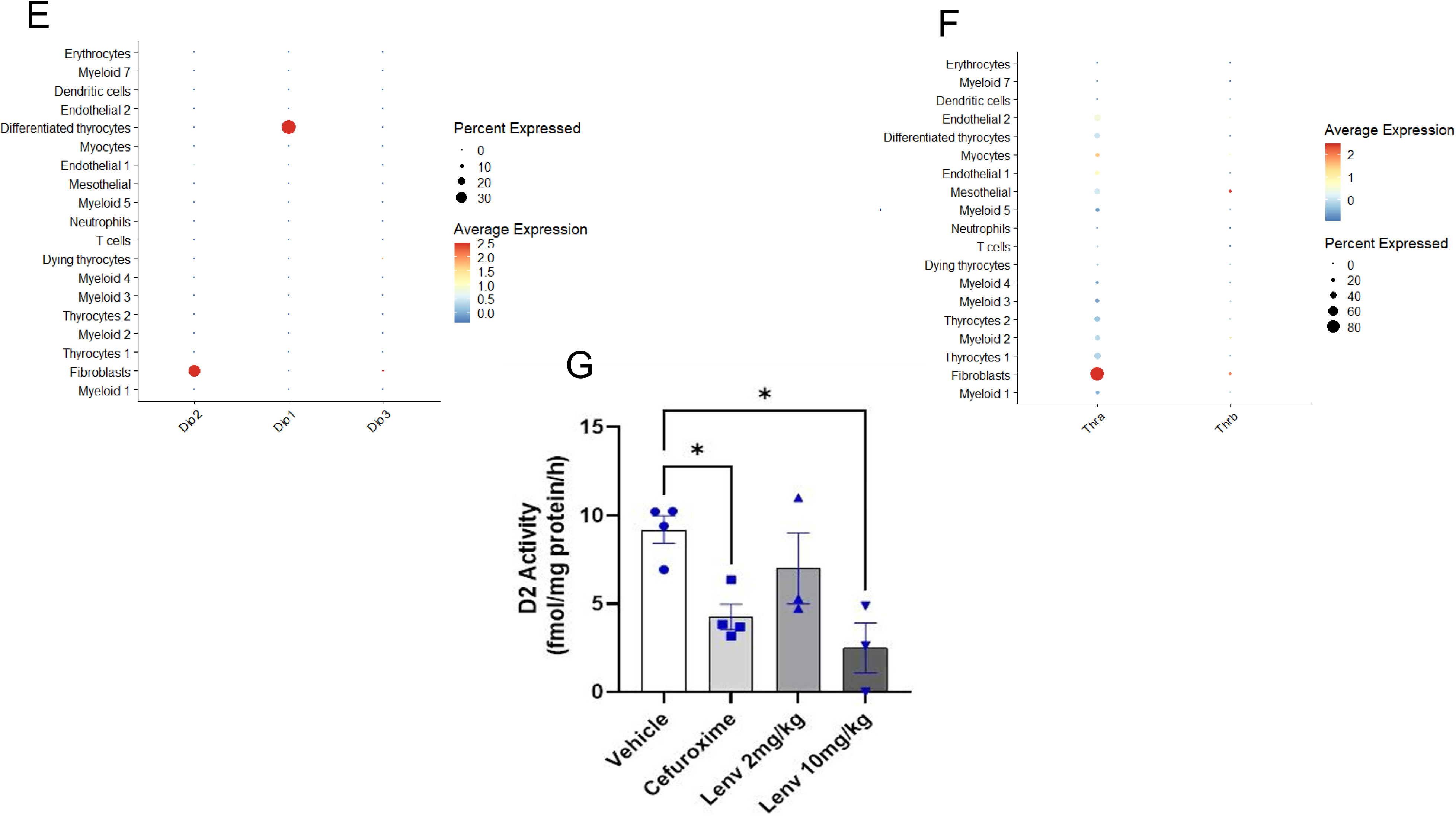
The TBP mouse model of DTC/ATC has *Dio2* expressed in CAF and D2 enzyme activity that is suppressed with cefuroxime. *TPO-CreERT2/LSL-BrafV600E/wt/Trp53*Δ*ex2-10/* Δ*ex2-10* (TBP) mice are treated with tamoxifen at 6-8 weeks of age and tumors are allowed to grow for 14-30 weeks. A) Ultrasounds images of a normal mouse thyroid (above) and large bilateral thyroid cancer in a TBP mouse approximately 20 weeks after tamoxifen induction of BrafV600E and loss of Tp53 (below). IHC was performed on TBP tumors for collagen 1A1 (Col1a1) to determine portion of fibrosis in these tumors. Images were scanned into the Aperio Digital Pathology system. After training the system, digital scanning produced area of positive staining (% pixels) and intensity of staining (1+, 2+ or 3+). B) Representative imaging from a large ATC tumor. C) Digital analysis results of whole tumor. Negative staining is predominantly tumor cells. D) UMAP features plot from scRNASeq of a female mouse harvested 20 weeks after tamoxifen injection (54 mg tumor) showing *Dio2* expression exclusively in the CAF. E) Dot Plot showing that *Dio2* was expressed in approximately 30% of the CAF. *Dio1* was expressed in approximately 30-40% of differentiated thyrocytes. *Dio3* was not expressed in any of the clusters. F) Dot Plot showing that *Tra* was expressed in approximately 80% of CAF, and much lower levels in the other cell clusters. *Trb* was expressed at very low levels in a small number of cells. G) TBP mice were treated with IP tamoxifen at 6-8 weeks old to induce *Brafv600e* and *Tp53* loss. At approximately 20 weeks of age, mice underwent thyroid US every 2 weeks. When tumors were 25-75 mm3, mice were randomized to vehicle (n=4) or cefuroxime (900 mg/kg/day) (n=4) daily IP injections for 15 days. At sacrifice, tumors were removed, snap frozen and then processed for D2 activity assays. D2 enzyme activity was measurable in all tumors. Cefuroxime inhibits D2 enzyme activity by approximately 50%. Low dose lenvatinib modestly inhibits D2 activity, while higher dose lenvatinib inhibits D2 activity by approximately 70%. * - p <0.05.

**Figure 14.**
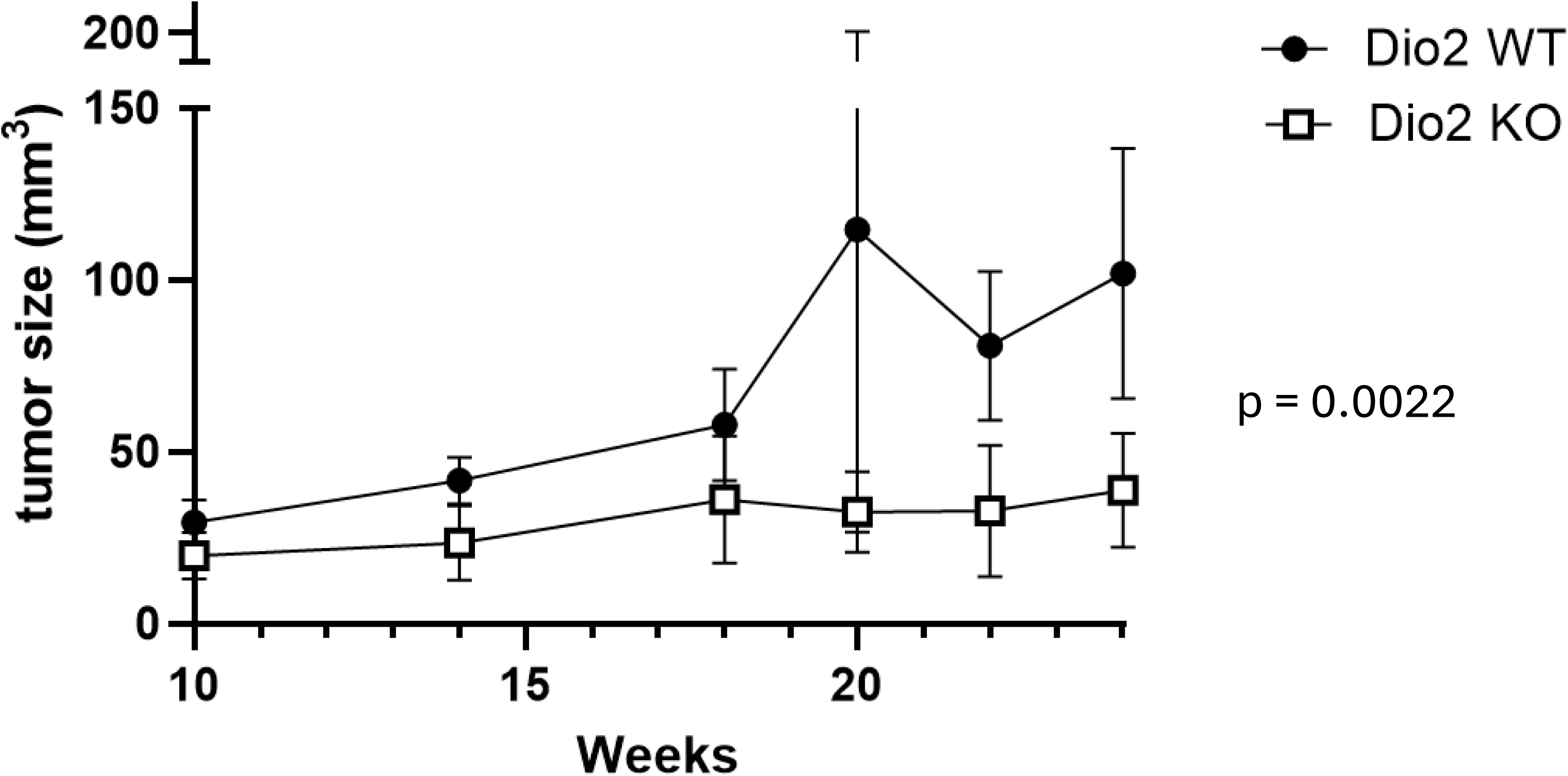
TBP mice lacking Dio2 (KO) have significantly slower tumor growth compared with TBP mice with intact Dio2 (WT). To genetically mimic complete D2 activity suppression in the TME, *Dio2* KO mice were back-crossed into TBP mice. Parental pairs of mice for the experimental mice were *TPO-CreER^T2^/Braf^V600E/V600E^/Trp53*^Δ*ex2–10/*Δ*ex2–10*^*/Dio2^+/-^*crossed with *TPO-CreER^T2^/Braf^WT/WT^/Trp53*^Δ*ex2–10/*Δ*ex2–10*^*/Dio2^+/-^*to generate littermates that were *TPO-CreER^T2^/Braf^V600E/WT^/Trp53*^Δ*ex2–10/*Δ*ex2–10*^ with WT or KO Dio2. Genetic alterations in Braf and Trp53 were induced by tamoxifen injection at approximately 8 weeks of age. After induction, mice were treated with 0.2 mg/L LT4 in the drinking water. Thyroid tumors were measured in 3 dimensions by ultrasound starting at 10 weeks after tamoxifen induction. Tumor growth was monitored by ultrasound over 14 weeks in TBP-Dio2WT (n= 19) and TBP-Dio2KO (n= 16) littermate mice.

### Working hypothesis

Based on the data that we and others have generated, we have developed a working hypothesis on the role of DIO2 in CAF and the TME (Figure 15). We propose that the primary effect of DIO2 is directly in the CAF to generate T3-signaling and affect the CAF phenotype to become a more tumor-promoting CAF, directly through CAF function and through CAF secretome effects on the other cells in the microenvironment. Furthermore, we propose that inhibition of DIO2 in CAF will reduce local T3 in the CAF, alter CAF function and create a less tumorigenic microenvironment.

**Figure 15.**
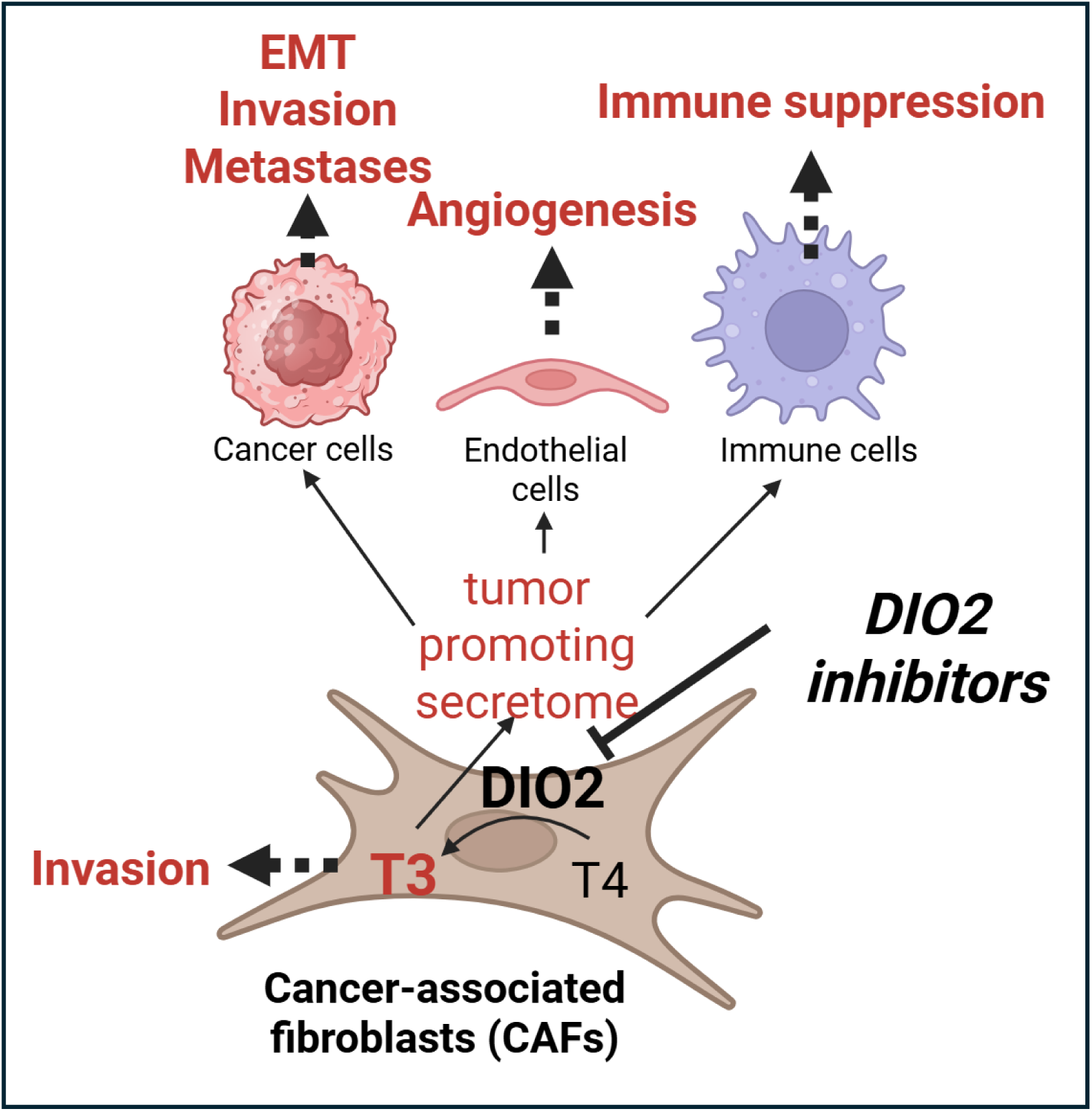
Working hypothesis on the role of DIO2 in cancer-associated fibroblasts (CAF) and the tumor microenvironment (TME). This figure was generated using BioRender.

## Discussion

We have identified DIO2 expression in CAF from multiple solid tumors. The tumor microenvironment (TME) plays a central role in cancer progression, particularly in aggressive subtypes such as anaplastic thyroid carcinoma (ATC). In this study, we identified *DIO2* expression in cancer-associated fibroblasts (CAF), with enrichment in myofibroblastic CAF (myCAF) across thyroid cancer and multiple other solid tumors. Using single-cell RNA sequencing (scRNA-seq) from a primary ATC tumor, we demonstrate that *DIO2* expression is restricted to CAF, with minimal or absent expression in malignant epithelial, immune, or endothelial populations. This finding was validated in a large thyroid cancer atlas dataset (21), where *DIO2* expression was most prominent in myCAF and to a lesser extent in inflammatory CAF (iCAF). The strong correlation between *DIO2* and canonical myCAF markers such as FAP, COL1A2, and POSTN in large bulk RNA-seq datasets further supports this association. Notably, the weaker correlation between *DIO2* and iCAF markers suggests that thyroid hormone activation may be preferentially linked to matrix-remodeling CAF functions rather than inflammatory signaling. These results extend prior observations that CAF are transcriptionally heterogeneous and functionally specialized (33, 34) and identify DIO2 as an important feature of a CAF subset.

De Stefano and colleagues recently published *DIO2* expression in thyroid cancer CAF using scRNASeq from publicly available data (10). They showed that *DIO2* expression was higher in the iCAF than the myCAF. Differences in our studies may be due to different definitions of myCAF and iCAF, and that the De Stefano group examined scRNASeq for only one of the seven studies (35) in the Thyroid Atlas we used for our studies. Furthermore, our larger bulk RNASeq data from the ORIEN database confirms the strongest association of *DIO2* expression and expression of myCAF markers. Nevertheless, both our studies independently showed that *DIO2* is expressed in thyroid cancer CAF, which is novel.

Our findings suggest that CAF are a critical site of local thyroid hormone activation within the TME, mediated by DIO2-dependent conversion of T4 to T3. The higher expression of *THRA* in CAF compared with other cell types further supports the concept of autocrine thyroid hormone signaling within stromal compartment. While thyroid hormone signaling has been implicated in cancer biology, including proliferation, metabolism, and angiogenesis (36), its spatial regulation within the TME has not been well defined. Our data indicate that CAF—not tumor cells—may be the dominant site of T3 generation in different cancers, suggesting a model in which stromal cells actively shape hormone-dependent signaling niches.

To our knowledge, the first description of deiodinase activity in fibroblasts was published by Refetoff and colleagues in 1972 (37). They showed that cultured human skin fibroblasts could convert T4 into T3, suggesting deiodinase activity in these fibroblasts. This same group later documented D2 activity in cultured skin fibroblasts (38). There is a fair amount of evidence that optimal wound healing requires thyroid hormone and local T3 in humans and mouse models (39–41). While there is some evidence for the importance of DIO3 in wound-healing (42, 43), the role of DIO2 is less clear. The phrase “tumors are wounds that do not heal” was attributed to Dr Harold Dvorak, who highlighted the similarities between tumor stroma generation and wound healing (44). We have shown that both *DIO2* and *DIO3* are expressed in many different tumor types (Figure 7), but that *DIO2* is expressed at much higher levels. We predict that both DIO2 and DIO3 may play an important role in wound healing and tumor stromal function, and that DIO2 may play a larger role in tumor stromal function and tumor aggressiveness.

We have used the unique ORIEN database to show that myCAF markers are associated with worse overall survival (OS) in many different cancer types (Figs 10 and 12), which is consistent with a recent metanalysis (45). Interestingly, one of the breast cancer studies showed an inverse survival relationship with myCAF markers, which we also found in our study using two different myCAF markers, *FAP* and *POSTN* (Fig 12). An older metanalysis also found a correlation between *FAP* expression and worse OS, particularly in pancreatic cancer (46). The recent Thyroid Atlas study showed a worse prognosis in thyroid cancer patients with higher myCAF scores (21). Our study provides independent confirmation that prognosis and survival across many different cancer types is associated with myCAF burden in those tumors. We also examined the relationship of the other major CAF subtype, iCAF, with OS using the ORIEN dataset. *CXCL12* expression was associated with worse OS in HNSCC, ovarian and pancreatic cancers (Fig 12), compared with a worse OS association with myCAF markers in 5 of the 7 tumor types tested. Furthermore, the HR for the iCAF association with OS in these tumors is lower compared with the myCAF markers, suggesting a weaker association. There is an ongoing debate about the tumor-promoting and restraining roles of myCAF and iCAF in solid tumors.

Our data suggest that myCAF may have a stronger role as tumor-promoting CAF in solid tumors. We found that the iCAF marker, *CXCL12*, was inversely associated with OS in breast cancer and melanoma, indicating that iCAF may play a more prominent role as tumor-restraining CAF in these cancers.

Our group and De Stefano (10) have shown that *DIO2* is expressed in CAF from thyroid cancers. We have also shown that *DIO2* is expressed in CAF from at least six other tumor types (Fig 7).

Our scRNASeq data further show that these CAF do not appear to express *DIO1* or *DIO3*. Our analysis of the bulk RNASeq data (ORIEN) shows a good correlation between *DIO2* and *FAP* (myCAF), supporting our scRNASeq data that *DIO2* is expressed in myCAF. Thyroid cancer shows an inverse correlation between *DIO2* and *FAP* expression using bulk RNA sequencing, likely due to the loss of *DIO2* expression in dedifferentiating thyroid cancers cells, outweighing expression in the CAF. *DIO3* has a weaker correlation with FAP expression across all tumor types, and *DIO1* shows no correlation with the myCAF marker, except for the expected inverse correlation in thyroid cancer. When we examined the relationship between *DIO2* expression and OS in these cancers, we found a significant correlation in pancreatic and bladder cancer, showing, for the first time, that there is a strong association of DIO2 expression and worse survival in cancer. We observed a significant negative association in breast and thyroid cancer. The inverse correlation may be related to the known expression of DIO2 in normal thyroid and breast tissue.

We have begun preclinical studies on the TBP (***T****PO-CreERT2/LSL-**B**rafV600E/wt/Tr**p**53*^Δ*ex2-10/*Δ^*^ex2-10^*) mouse model. This is a transgenic model that develops PTC and ATC (25). We show here that these tumors have extensive fibrosis, suggesting that this is a good model to study the role of CAF in tumor progression. Jolly and colleagues have previously shown that a different *BrafV600E*-driven model of thyroid cancer also has extensive fibrosis (47), where as a RAS-driven model of thyroid cancer has much less fibrosis (48). Our scRNASeq analysis of the TBP thyroid cancer shows that *Dio2* is exclusively expressed in the CAF and *Thra* is highly expressed in the CAF as well, making this an excellent model to study the role of Dio2 in the TME. Treating the TBP mice with cefuroxime, a known inhibitor of D2 activity, showed an approximately 50% reduction in tumor tissue D2 activity. Interestingly, lenvatinib, the most commonly used MKI in advanced thyroid cancer, reduced D2 activity by approximately 70% in a dose-dependent manner. Others have shown that the kinase inhibitors selpercatinib and vandetanib reduced D2 activity (30, 31), but we are the first to show that lenvatinib inhibits D2 activity. This may be part of the mechanism of action of lenvatinib in advanced cancer, which can be explored in our TBP mouse model. Finally, we have used the TBP model of papillary and anaplastic thyroid cancer to begin to understand the role of Dio2 in the TME. We showed that TBP mice lacking *Dio2* had slower tumor growth compared with TBP mice with normal levels of *Dio2*, which indicates that Dio2 is critical in the TME, and could be a good target for inhibition across multiple types of cancer.

We have developed a working hypothesis on the role of DIO2 in CAF and TME (Figure 15). We propose that the primary effect of DIO2 is directly in the CAF to generate T3-signaling and affect the CAF phenotype to become a more tumor-promoting CAF, directly through CAF function and through CAF secretome effects on the other cells in the microenvironment. An alternative hypothesis is that DIO2 not only provides T3-signaling in the CAF but also provides T3 for increased thyroid hormone action in other cells of the TME, which is testable. While our scRNASeq analysis shows that *DIO2* expression is limited to CAF in the TME across multiple tumor types, other groups have shown that *DIO2* is also expressed in the epithelial-derived cancer cells in thyroid cancer (10, 49, 50) and that this expression may be TP53 dependent (51). Interestingly, De Stefano and colleagues recently showed that a human thyroid cancer cell line co-cultured with mouse mesenchymal fibroadipogenic progenitor (FAP) cells lacking *Dio2*, had increased *DIO2* expression (10). This would suggest that DIO2 is critical in the TME and can be expressed either in CAF or epithelial-derived cancer cells. It is not clear if this also occurs in other cancers.

In summary, we have shown that DIO2 is a conserved marker of CAF, predominately myCAF, across multiple solid tumors and is associated with adverse clinical outcomes in some cancers. The TBP mouse provides a robust preclinical model for defining the mechanistic role of DIO2 in CAF biology and evaluating DIO2-directed therapeutic strategies targeting the tumor microenvironment.

## Data Availability

All data produced in the present study are available upon reasonable request to the authors

## Acknowledgements

We thank Dr Alice Batistuzzo for her help with the D2 assay. This work was supported by a pilot grant from the Colorado HNC SPORE grant (P50 CA261605) to BRH, an NIH F30 CA281125 grant and NIH T32 GM007347 grant to MAL, NIH VCORCDP K12CA090625, K08CA240901, and R01CA272875 grants to VLW, and by a pilot grant from the School of Medicine at the University of Colorado Anschutz Medical Campus. This work was also supported by a generous gift from Gloria and Michael Komppa, and the Mary Rossick Kern and Jerome H Kern Endowment.

## Conflicts of Interest

BRH is Clinical Liaison for Sonic Healthcare USA, a molecular diagnostics company.

VLW is a co-founder of PalinGenix Therapeutics, a company focused on the development of drugs targeting developmental signaling pathways.

## Notes

### Author Declarations

Approval for analysis of human ATC tissue used in single cell sequencing was approved by the Colorado Multiple Institutional Review Board (COMIRB) Approval for use of human data in the Oncology Research Information Exchange Network (ORIEN) database was approved by ORIEN

